# Bit-Exact Reconstruction and Certification of Electrocardiogram Waveforms from Vector-Encoded PDF Files

**DOI:** 10.64898/2026.09.23.26362281

**Authors:** Khaled Abdelrahman, Hans Friedrich Stabenau, Konstantinos Patlatzoglou, Arunashis Sau, Boroumand Zeidaabadi, Libor Pastika, Nicholas S. Peters, Fu Siong Ng, Daniel B. Kramer, Jonathan W. Waks

## Abstract

ECG waveform data, which is increasingly required for artificial intelligence analyses, is often unavailable in raw data formats. Digitization can approximate original raw data from ECG images, but digitizers reconstruct ECG waveforms only approximately. Exact waveform reconstruction is needed to guarantee that any errors, even if small, do not unexpectedly propagate into downstream analyses. Portable document format (PDF) files containing vector graphics store waveforms derived directly from raw data. This improves reconstruction accuracy, but existing PDF reconstruction methods remain approximate because raw data values are rounded during PDF encoding/generation. Despite rounding, however, there is a 1:1 mapping between raw data and PDF-encoded values which can be reversed to recover the original raw data with no error. We developed open-source software in MATLAB and Python (http://github.com/BIVectors/pdf2ECG) for bit-exact waveform reconstruction of General Electric MUSE ECG PDFs based on double quantization of ECG waveform data. The software supports multiple PDF layouts and voltage gains, automatically flags waveform clipping under which exact reconstruction is not possible, and certifies if parameter values used for waveform reconstruction are consistent with parameter values used during PDF encoding. Accuracy was assessed by comparing PDF reconstructed waveforms to raw XML waveforms, and reconstruction parameters were perturbed to determine the accuracy of waveform certification. 1,632 PDF-XML pairs were assessed in 5 layouts, each at 3 gains. After software-identified waveform clipping was excluded, the software reconstructed 100% of ECG waveforms with zero error compared to ground-truth XML. Certification tests were extremely sensitive to minor perturbation in encoding parameters, and if passed, confirmed bit-exact waveform reconstruction without need for ground-truth comparisons. Although this method does not replace the need for general ECG image digitization, for MUSE generated PDFs this method is preferable to rasterization followed by image-based digitization.

## 1. Introduction

In clinical practice, electrocardiograms (ECGs) are usually reviewed and interpreted from images, either on a computer screen or printed on paper. Recent advances in artificial intelligence ECG analysis, and other more conventional ECG signal processing analyses such as vectorcardiography, have made obtaining raw, digital ECG waveforms increasingly important. Raw waveforms are typically exported from ECG repositories in formats such as Extensible Markup Language (XML), Digital Imaging and Communications in Medicine (DICOM), or plain text, where ECG lead voltage over time is encoded as numeric data that can be read and further processed by a computer program.

However, although raw ECG waveform data are increasingly in demand, exporting raw waveforms can be a logistical challenge, and often requires special access to ECG systems. Additionally, some ECGs do not exist in a digital database which allows export of raw waveforms, and images may be the only ECG data that are available. Many clinical studies collect and save ECG images, often in portable document format (PDF), rather than as raw waveform data, because images (and not the raw numeric data) are what is interpreted by physicians, and image files are easier to review, process, and store. There is, therefore, significant interest in improving the ability to digitize ECG images to avoid having to rely on raw digital waveform files which can be difficult or impossible to obtain in some situations. As a result, there are now many algorithms and software programs that take an image of an ECG, and through signal processing and/or machine learning, approximate the raw ECG waveforms so they can be used for downstream computational tasks [1–8].

Although these digitization programs perform well overall, there remain limitations in how accurate such programs can be, as they have to account for noise and variable image quality, variable alignment and orientation, ambiguity in zero voltage references, extraction and conversion between pixels and physical units, overlapping signals, and the fact that the lines used to draw the ECG waveforms have width and therefore represent a range of values rather than a single value at each time point. As a result, even “perfect” quality images will have some digitization error, and state-of-the-art ECG image digitizers therefore tend to have root-mean square errors (RMSE) on the order of 20 − 100 *μ*V [1–8], with variation based on image quality and different digitization algorithms.

An underused property of ECG images stored in PDF format is that they can utilize vector graphics, where the displayed waveforms are stored as a series of points which are derived directly from the raw ECG waveform data, rather than rasterized images. These PDFs therefore contain information on the actual voltage values that would be present in raw digital formats such as XML, but they have the benefit of being more readily available, and exporting these files usually does not require any special access. Vector extraction also avoids rasterization-related image quality loss and line width and overlap ambiguity because each waveform remains a separate, well-defined, graphics object.

Several groups have reconstructed ECG waveform data from vector-encoded PDFs [9–12] although most assessed fidelity by visual comparison or downstream use rather than assessment of quantitative error of multiple PDF display formats and gains against ground-truth waveforms. Shie et al. [9] recently reported results from ECG waveform reconstruction in a large dataset of vector PDF ECGs in a single layout format and 10 mm/mV gain with very low mean absolute error (MAE) of ~0.26 *μ*V measured against system-exported raw waveforms. Although the errors associated with PDF vector data extraction are, not surprisingly, significantly lower than when digitizing rasterized images, residual errors remain because the PDF rendering engine rounds fractional waveform amplitudes to integer stream coordinates which are stored within the PDF and then plotted. Any method that reconstructs physical units from the PDF stream coordinates by directly converting stream units into mV, therefore carries a minimum error of ±0.5 stream units, with an expected mean absolute value of ~0.25 stream units (~0.25 *μ*V at 10 mm/mV), closely matching previously reported values [9]. This error is inversely related to display gain, and is twice as large at 5 mm/mV.

Although it would seem that rounding performed during the conversion between raw ECG data and PDF vector stream units destroys the original unrounded data, because the PDF stream unit grid is finer than the raw analogue-to-digital unit (ADU) grid, this rounding is not information-destroying. Each waveform encoded in the PDF has been quantized twice [13, 14], distinct ADU values map to distinct stream values, and the structure left by the second quantization permits *exact* recovery of the original ADU integers rather than an approximation of them. Therefore, it is possible to perform *bit-exact* waveform recovery, where the recovered waveform values are *exactly* the same as the original raw data which were used to generate the PDF (as would be obtained in an equivalent XML file exported directly from the ECG database), rather than a close approximation.

For many ECG analyses, such as measuring heart rate or interval measurements, residual sub-*μ*V errors are unlikely to be significant. The error, due to rounding, however, is not completely negligible. Because the rounding error is not independent and random noise and is a function of signal amplitude, in certain downstream uses it could introduce significant variability in results between PDF reconstructed and directly-exported raw data. A very low MAE or RMSE in waveform reconstruction therefore does not necessarily guarantee a similarly low level of error for *all* downstream uses. As a result, an approximate reconstruction, even if extremely accurate, cannot be guaranteed safe for all potential downstream uses, and each new analysis must independently establish that the residual error does not matter for it specifically. Exactness also makes waveform reconstruction certifiable. PDF stream units and the reconstructed ADUs have a structure determined entirely by the encoding, and as a result, a waveform reconstruction can be internally certified under the PDF encoding model without requiring reference to raw waveforms. Such certification of bit-exactness is not possible with an approximate reconstruction, regardless of accuracy.

A second, distinct source of error, which can affect any PDF or image digitization pipeline, arises from signal “clipping”, where portions of the signal exceeding the bounds of the plotting area are not displayed and are therefore completely absent from the displayed output. Unlike the rounding errors described above, clipping destroys information. No reconstruction method, approximate or exact, can recover part of a waveform that was never displayed, and the resulting error cannot be determined from the PDF alone. Despite its importance to ECG image digitization, the prevalence and downstream relevance of clipping have not been well characterized. Clipping, however, is detectable from the PDF itself, so a recovered waveform can be explicitly flagged as incomplete.

Existing programs, which perform approximate PDF waveform recovery, also only support a single PDF layout and gain [9–11], limiting practical use on real-world archives that contain mixed formats/gains. To our knowledge, no existing software performs bit-exact PDF waveform reconstruction or provides any means to certify that a given reconstruction is correct. The purpose of this manuscript was therefore to develop and validate open-source software (in both MATLAB and Python) to reconstruct bit-exact ECG waveforms from General Electric (GE) MUSE format ECG PDFs, as recovering the original waveform exactly overcomes a significant limitation: when the reconstructed waveform is identical to the source, every downstream result is also guaranteed identical to that obtained from the original data, without the need for per-use validation. The method operates across multiple MUSE PDF display layouts and voltage gains, and detects and flags waveforms with signal clipping where reconstruction cannot be bit-exact. It also returns a ground-truth-free certification that the reconstructed waveforms satisfy the encoding parameters required for bit-exact recovery to ensure that any PDFs with unexpected parameters that would prevent bit-exact waveform reconstruction are not silently propagated into downstream analyses.

## 2. Methods/Software Description

### 2.1. Study Population

A database of sequential 12-lead ECGs performed at Beth Israel Deaconess Medical Center (BIDMC, Boston, MA) were selected for inclusion in this study. These ECGs were extracted from the GE MUSE client version 9.0.7 (in 2021) and version 10.2.3 (in 2026) in different formats including XML (which contains the raw signals in base64 encoded integer ADUs with 4.88 *μ*V per ADU), and single page PDF files with 12 10-second rhythm strips (12×1 format), 3×4 2.5-second leads with 0 (3×4+0 format), 1 (3×4+1 format) or 3 (3×4+3 format) 10-second rhythm strips, and 6×2 5-second tracings (6×2 format). All ECGs were acquired at 500 Hz. Using PDF and XML metadata, the different formats were linked to each other with unique identifiers. There were no restrictions on ECG quality or diagnosis, and multiple ECGs per patient were also allowed. ECGs were excluded from the study only if there were different filtering settings used in the PDF and XML. The study was approved by the BIDMC Institutional Review Board (2003P000420).

### 2.2. PDF Waveform Reconstruction Algorithm Overview

The PDF waveform reconstruction algorithm, as summarized in Figure 1, was designed with the goal of extracting the coordinates of polylines that are used to render the ECG waveforms, and then utilizing the double quantization of the PDF datastream to convert back to the original bit-identical ADUs which are stored in the equivalent XML file. A summary of variables used in the subsequent sections is available in Supplemental Table 1. Software was developed in MATLAB R2022a (MathWorks, Natick, MA) and ported to Python 3 (Python Software Foundation; Python 3.14.3 with numpy 2.4.5). Source code is available at http://github.com/BIVectors/pdf2ecg under GNU General Public License version 3.

**Figure 1:**
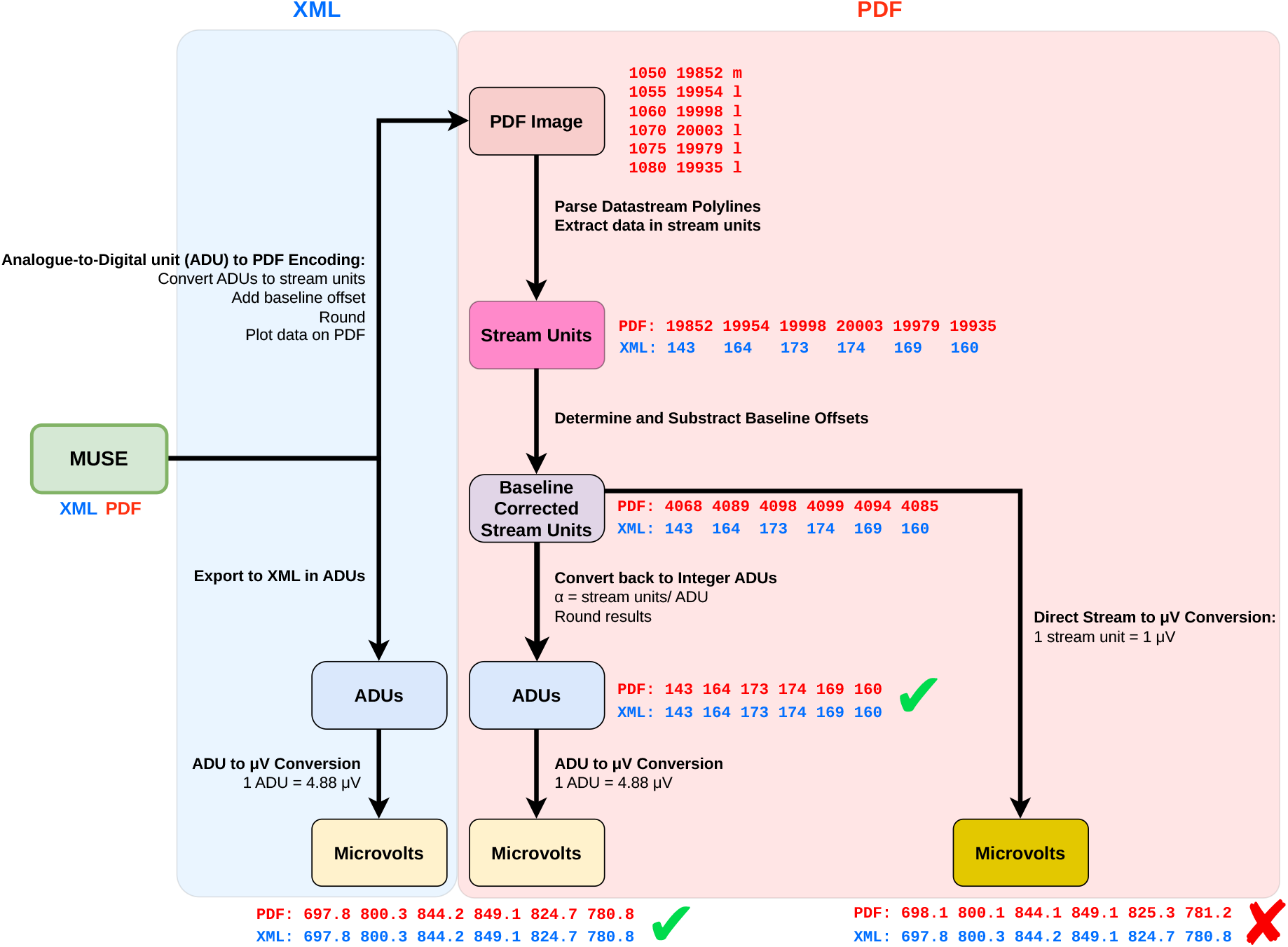
Summary of the ECG waveform reconstruction algorithm. MUSE can output ECGs as raw data in XML format, where data is expressed in analog to digital units (ADUs) (1 ADU = 4.88 *μ*V), or as vector-encoded PDF images. The blue shaded area and 6 blue waveform values denote the XML extraction pathway and XML ADUs which are then converted to physical units (*μ*V). The red shaded area and 6 red waveform values denote various steps in the PDF reconstruction algorithm, which is explained in detail in the Methods. For PDF reconstruction, data is first extracted from the PDF data stream into integer stream units. Baseline offsets are subtracted, and then the stream:ADU ratio, *α*, is used to convert back to ADUs prior to rounding. If stream units are converted directly to physical units such as mV by simply subtracting the zero reference and dividing by the stream-to-mV conversion factor (right-most pathway), there will always be a small residual error in the recovered waveform due to rounding used when ADUs are converted to stream units at the time of PDF generation. Reconstructed ADUs have bit-exact agreement with the ADUs directly exported from MUSE in XML format, and can be multiplied by 4.88 *μ*V/ADU to obtain physical units.

### 2.3. PDF Data Structure

PDF files are processed using published specifications [15]. Raw data from the PDF file is read as unsigned 8-bit integers. Although some header information can be read directly from this 8-bit data, the actual ECG signals and other metadata are encoded in data “streams” which are data objects that include a dictionary followed by data bracketed between the keywords “stream” and “endstream”. The length of each stream is determined from the direct `/Length` entry in the stream dictionary rather than searching for an “endstream” marker to avoid errors when binary compressed bytes happen to contain the ASCII pattern “endstream”. MUSE PDFs all use the zlib deflate compression algorithm for stream data, which is indicated by the dictionary entry `/FlateDecode`. In general, all tested MUSE PDFs have a single stream per page, although the software can handle multiple streams.

### 2.4. Parsing the Decompressed Stream and Extracting Polylines

PDF data streams use a stack-based language which utilizes numeric operands and alphabetic operators [15]. The decompressed content stream is tokenized on whitespace and the resulting sequence is interpreted with a stack-based PDF path operator program. Numeric operand tokens are consumed by alphabetic operator tokens to generate polyline graphics objects. For the tested MUSE PDF files, 3 operator types are required to reconstruct waveform coordinates/metadata: “m” (moveto) which starts a new polyline, “l” (lineto) which appends a coordinate to the current polyline, and “S” (stroke) which ends and stores the polyline. Additional operators “RG” and “G” which set the color of polylines, and “cm” which loads the affine transformation matrix used to map stream coordinates to physical units can be processed, although the current version of the software does not utilize them. Other encountered operators were confirmed not to affect the waveform paths used by the algorithm. An example of parsing data into polyline objects is shown in Supplemental Section S.1.

A typical PDF will have hundreds of polylines corresponding to the ECG lead waveforms, ECG grid lines, calibration signals, and other PDF decorations. Each of these polylines has a distinct set of features allowing for their identification. The horizontal or vertical minor gridlines are identified as polyline objects that consist of 2 points with the same *x*- or *y*-coordinate, respectively, spanning at least 50% of the page to filter out marks for changing leads. Calibration signals are identified as polyline objects that contain 60 points and only 2 voltage values. To isolate the ECG waveforms, a length filter excludes any polyline that is ≤ 500 samples in length (this allows some variation for lower/higher sampling frequencies that might be encountered). The distribution of polyline lengths and their identification is shown in Supplemental Figure 1. After this length filtering, the remaining polylines (12-15 depending on the PDF layout) are the ECG lead waveforms. The ECG leads are then sorted on the *x*-coordinate of their first sample and within each column by the descending median *y*-coordinate, producing the standard clinical lead order of I, II, III, aVR, aVL, aVF, V1-V6. The lead order is verified by extracting lead names and starting coordinates out of the data stream, and any deviations from the expected lead layout result in an error. Any 10-second rhythm strips in the 3×4 format are identified by their labeling.

### 2.5. Determining ECG Gain and Sweep Speed

The decompressed stream is searched for information on the time and voltage calibration by looking for the characters “25 mm/s” and *G* “mm/mV” where *G* is an integer extracted by regular expressions. Gains of 5 mm/mV, 10 mm/mV, and 20 mm/mV are supported. Sweep speeds other than *T* = 25 mm/s are extremely rare, are not currently supported, and if present result in an error. PDFs with mixed limb-and-precordial gains are also not currently supported.

### 2.6. The Relationship Between Stream Units and Physical Units

We use data contained within each PDF file to determine the relationship between PDF stream units, ADUs, and physical units (mV or seconds). We start by building upon previous work [9, 10] where a polyline of known voltage amplitude is used to determine the relationship between stream units and physical units. Prior works used the calibration signal, which is a square wave present on every ECG that spans 1 mV of relative signal amplitude. At a standard gain of 10 mm/mV, each calibration signal spans 1000 stream units, indicating that 1 stream unit = 1 *μ*V. This relationship is also present at a gain of 5 mm/mV where each calibration signal spans 500 stream units, indicating that 1 stream unit = 2 *μ*V. However, this relationship breaks down at gains of 20 mm/mV due to the fact that the calibration signal in PDF plotted stream units is a rounded representation of the original 1 mV calibration signal waveform. On all PDF formats except 12×1, the 20 mm/mV calibration signal spans 2001 stream units (not 2000 as expected), which would suggest that 1 stream unit ≈ 0.49975 *μ*V, although this is incorrect. This discrepancy occurs because MUSE needs to render a 1 mV calibration signal, but it can only draw waveforms that are integer multiples of 1 ADU. Because 1 ADU = 4.88 *μ*V, the possible stream unit values are therefore constrained. There are approximately 204.92 ADUs in 1 mV, which rounds to 205 ADU. Therefore the true amplitude of the “1 mV” calibration pulse is actually 1.0004 mV. This excess 0.4 *μ*V is imperceptible, and therefore for plotting the ECG on paper it is not an issue. At 5 and 10 mm/mV, the excess signal does not cross 0.5 stream units, so rounding erases it, but at 20 mm/mV the excess signal does exceed 0.5 stream units, which results in 1 extra stream unit appearing in the calibration pulse. As a result, using the calibration pulse to dynamically assess the stream unit to mV relationship in each ECG will fail at gains of 20 mm/mV, requiring either hard coding the value (which is not ideal as the stream to mV relationship could change), or utilizing other features of the PDF that do not have this rounding limitation.

Instead of using the calibration signal, we utilize the ECG grid lines which are geometrically defined and always spaced 1 mm apart. Use of the grid lines therefore avoids having to account for the ADU grid and the relationship between ADUs and mV which may not be known up front and which involves rounding. The gain, which is easily read off the PDF (see Section 2.5 above), defines the relationship between mm and mV or seconds, and by measuring the spacing on the minor grid lines in stream units/mm and multiplying by the gain (in mm/mV or mm/s), we obtain the true relationship between stream units and mV and stream units and seconds. If we define the voltage minor grid spacing in stream units/mm as *g*_*v*_, the time minor grid spacing as *g*_*t*_ stream units/mm, the voltage gain as *G* mm/mV, and the horizontal sweep speed as *T* mm/sec, (which for the current software is fixed so at *T* = 25 mm/s), we obtain the relationship between stream units and mV (*b*_*v*_) or seconds (*b*_*t*_):

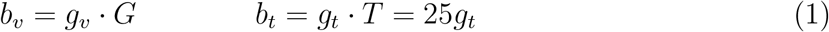

Nominally *g*_*v*_ = *g*_*t*_ = 100 stream units/mm.

A third option for determining the relationship between stream units and mV or seconds is to go sequentially through the PDF provided conversions from stream units to PDF points to mm and then to mV. The details of these conversions are provided in Supplemental Section S.2. We have provided information on this additional option in Supplement Section S.2 because, although it would seem that this approach, which uses conversion factors directly encoded within the PDF file, would be preferable to using drawn polylines, it has one subtle but critical flaw: the cm matrix encoded in the data stream, which provides the conversion from stream units to PDF points, is truncated at 5 decimal places. This degree of precision is fine for plotting the stream units on the PDF page, but errors on the order of 1 × 10^−5^ due to this truncation can propagate and cause rare and silent failures of waveform reconstruction (see Supplemental Sections S.2 and S.6 for additional information).

### 2.7. Determining the Baseline Offset

The baseline offset *B*, which is used to position each ECG waveform on the PDF page, must be known accurately to achieve bit-exact ADU reconstruction. Further information on how exact *B* needs to be known is available in Supplement Section S.7.

The software determines the value of *B* primarily by using the calibration signals which are extracted from polyline objects (see Section 2.4). Although the calibration signals are not an optimal choice for determining the stream unit-to-mV conversion for reasons discussed previously, their baseline, which by definition is set to 0 mV, can be used directly in stream units without any concerns about rounding. In all PDF layout formats except 12 ×1 there is a calibration signal on each row of ECG waveforms (including rhythm strips if present). We therefore determine the number of calibration pulses and their baselines in stream units. These values are used for *B* in each row, with the ECG leads that are in each row determined by the PDF layout. 12×1 format, however, only contains a single calibration pulse aligned with lead I. To determine the locations of the baselines for the remaining 11 leads in 12×1 format, we determine the spacing between leads by evaluating the stream unit coordinates of the ECG lead labels. Although the labels are not at the correct zero baseline values, the difference in stream units between labels is the same as the difference in stream units between calibration signals. We therefore determine the value of *B* for lead I, determine the difference between labels, and apply that difference to *B* in lead I to generate the *B* values for the remaining 11 leads. As shown below in Section 2.15 and Supplement Section S.5, we are able to verify that these values of *B* are correct by comparing values of processed PDFs to ground truth XML, and through certification tests that do not require any ground truth signals.

### 2.8. Directly Transforming Stream Units into Physical Units

As has been done by other groups [9, 10], converting stream units (*S*) to physical units 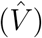 can be accomplished by subtracting the offset used to zero and position the waveform on the PDF page (*B*) and dividing by the stream unit-to-mV conversion factor *b*_*v*_ determined based on the grid spacing and gain as noted above in Eq. 1.

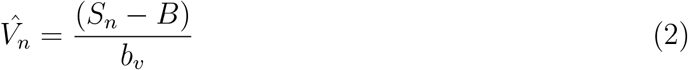

This conversion yields a low error when compared against the XML ground truth [9] (see Results Section 3.3). This error, like the error in the calibration pulses, arises from rounding introduced when integer ADU values (*A*) are converted to integer stream units (*S*) during PDF encoding, the baseline offset used to position the ECG lead on the PDF page (*B*), and the non-integer relationship (*α*) between stream units and ADUs.

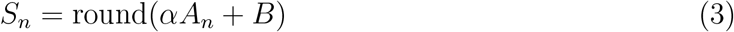

As a result of this rounding, there will be up to ±0.5 stream unit errors during the conversion from ADUs to stream units, and converting these rounded stream units directly to physical units will therefore never result in perfect signal reconstruction. Alternatively, it would be better to reverse the stream to ADU conversion, directly reconstruct the ADUs encoded in MUSE/the XML, and then convert these ADUs to physical units. Such a reverse transformation is non-trivial, as the rounding performed at the time of converting ADUs into stream units would seemingly “destroy” fractional stream units. However, we can exploit features of the ADU to stream conversion at the time of PDF generation to reverse PDF-encoded stream units back to ADUs. To do this we first need to determine the relationship between stream units and ADUs (*α*).

### 2.9. Determining the Stream Unit to ADU Relationship, *α*

The most important value we will use to reconstruct the bit-exact ADU values is *α*, which is the ratio of stream units to 1 ADU. The PDF grid line spacing (*g*_*v*_ in units/mm), PDF gain printed on the PDF (*G* in mm/mV), and the MUSE-specified voltage per ADU (*m*), which is nominally 0.00488 mV/ADU, provide the value of *α*. Combining *m* with Eq 1, we obtain *α*:

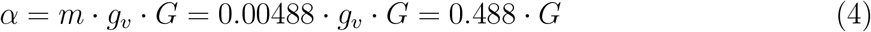

The PDF does not directly contain the value of *m*, but as we will show, we can use the PDF data alone to certify if the value of *m* used to derive *α* is correct (see Section 2.15).

### 2.10. Converting Stream Units Back to ADU Values

#### 2.10.1. Rationale for Converting Stream Units Back to ADUs

Although the errors associated with direct stream to mV conversion (Section 2.8 and Eq. 2) may be small in magnitude, it is not possible to guarantee that they will never propagate through downstream analyses and cause measurable differences when comparing use of the PDF-reconstructed vs. raw XML-derived waveforms. However, we can utilize the fact that each waveform has been quantized twice: first to integer ADUs by the ECG machine at the time of recording, and again to integer stream units at the time of PDF generation. Critically, the second quantization is *not* information destroying. Because the stream grid is finer than the ADU grid, distinct ADU values map to distinct stream unit values, and this encoding can be reversed. Recovering the exact ADU values that generated the PDF stream units is therefore not a problem of improving the accuracy of signal approximation, but is instead a problem of reversing a known quantization [13, 14].

#### 2.10.2. Evidence that ADU Values Can be Exactly Reconstructed

Direct evidence that exact ADU reconstruction is achievable appears in the structure of the difference between subsequent stream unit samples, Δ*S* = *S*_*n*+1_ − *S*_*n*_. Figure 2 shows the distribution of Δ*S* pooled across all 12 leads for 30 randomly selected ECGs. Stream units are always integers, and the deltas between samples occupy only a limited set of integers defined by:

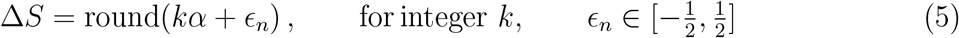

where *ϵ*_*n*_ is the error introduced by rounding. For a full derivation see Supplemental Section S.3. From Eq. 4, at a gain of 10 mm/mV, *α* = 4.88, and we therefore get the rounding intervals defined by 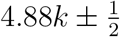 (see Figures 3 and 4):

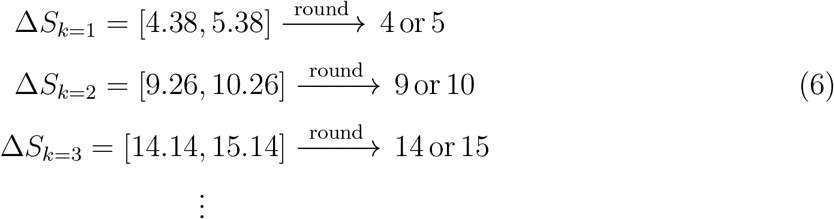

**Figure 2:**
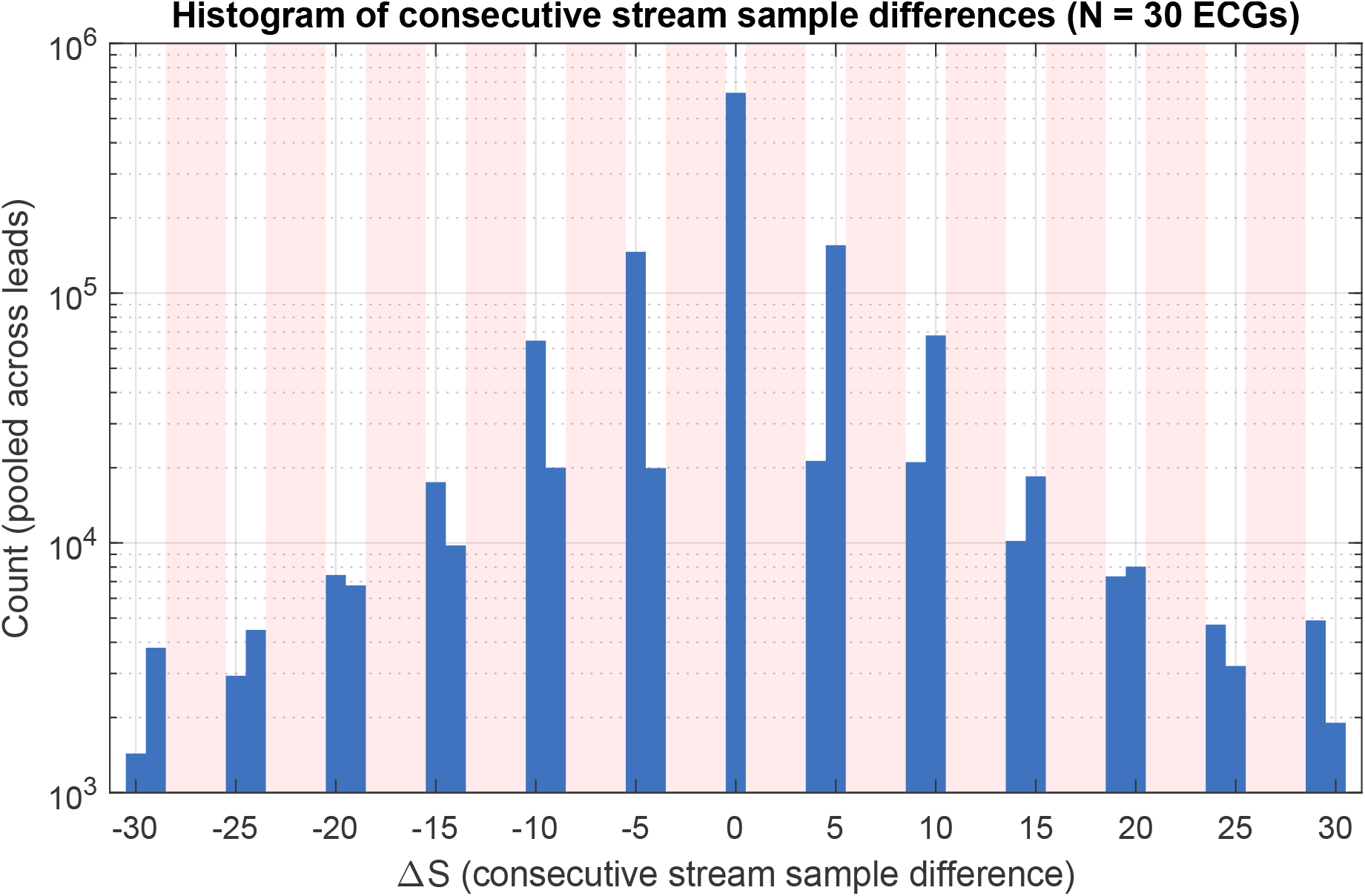
Histogram of the differences between consecutive stream samples Δ*S* = *S*_*n*+1_ – *S*_*n*_ for a voltage gain of 10 mm/mV. Blue bars indicate the number of times that a specific value of Δ*S* was observed across 30 ECGs x 12 leads displayed at log scale. Red shaded areas indicate bins where 0 counts are observed. Because the ratio of quantization for stream units and ADUs for a gain of 10 mm/mV is *α* = 4.88, Δ*S* can only take on values of {0, ±4, ±5, ±9, ±10, ±14, ±15, …}, which correspond to the integers bracketing *αk* for each integer ADU step *k*. The values {±1, ±2, ±3, ±6, ±7, ±8, ±11, ±12, ±13, …} are never observed. This pattern is consistent with integer rounding applied to a signal with *α*-quantization. It also confirms that the encoder from ADU values to stream units did not introduce significant smoothing, dithering, or filtering. For unclipped samples, this one-to-one mapping preserves the information needed to invert the stream encoding when the scale and baseline parameters are correctly identified. Further information is available in the Methods and Figures 3 and 4.

**Figure 3:**
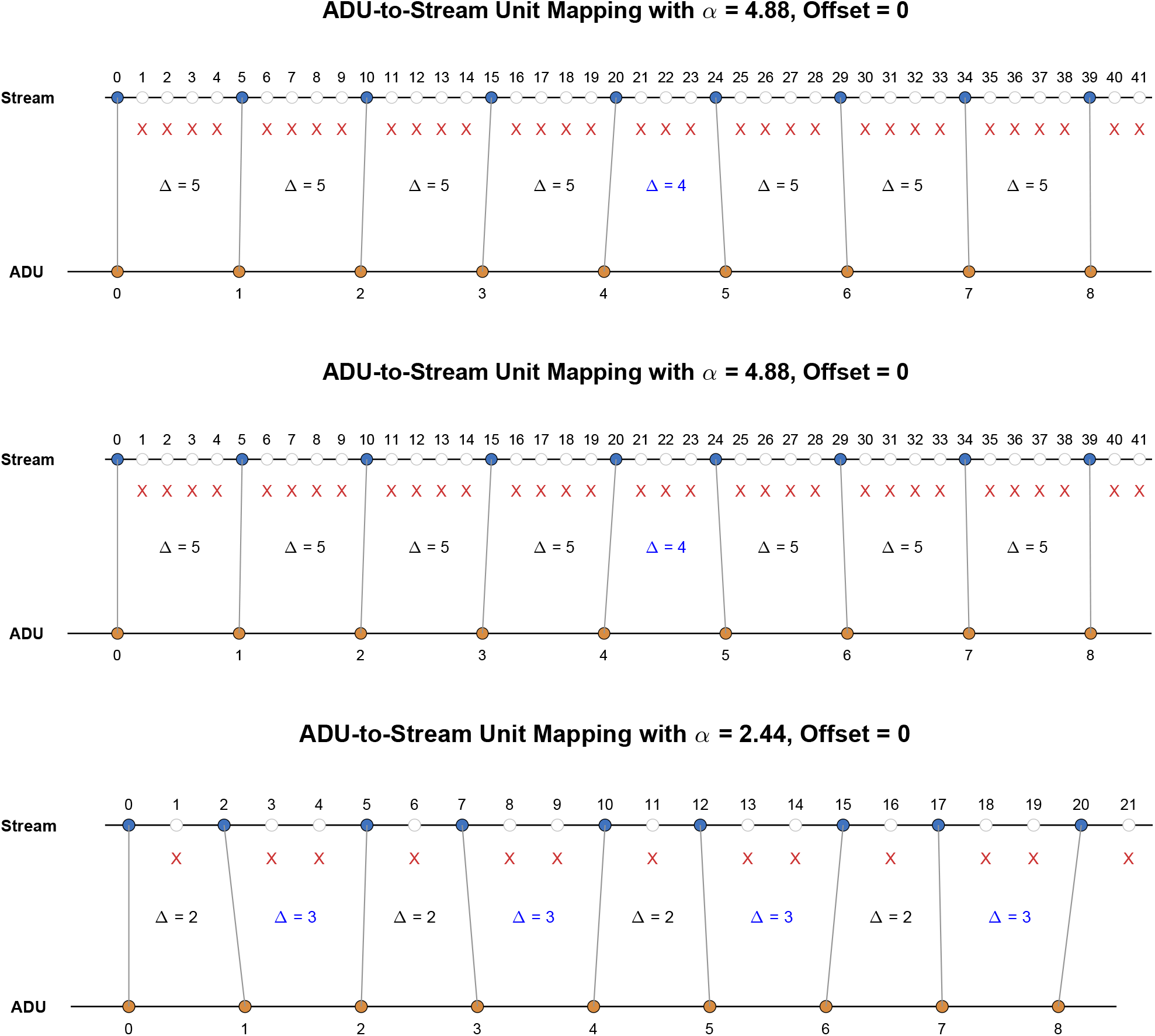
Conceptual illustration of double quantization when mapping integer ADU values to integer PDF stream units using different values of *α* and different offsets. As the ADU values are monotonically increasing, this is equivalent to a 1 ADU Δ*S*_1_. Note that there is always a unique mapping between ADUs and stream units. **Top and middle Panels**: When *α* = 4.88, the difference between stream units is always either 4 or 5 (integers that span *α* = 4.88) for a 1 unit change in ADU, regardless of the offset. **Bottom Panel**: When *α* = 2.44, the difference between stream units is always either 2 or 3 (integers that span *α* = 2.44)

**Figure 4:**
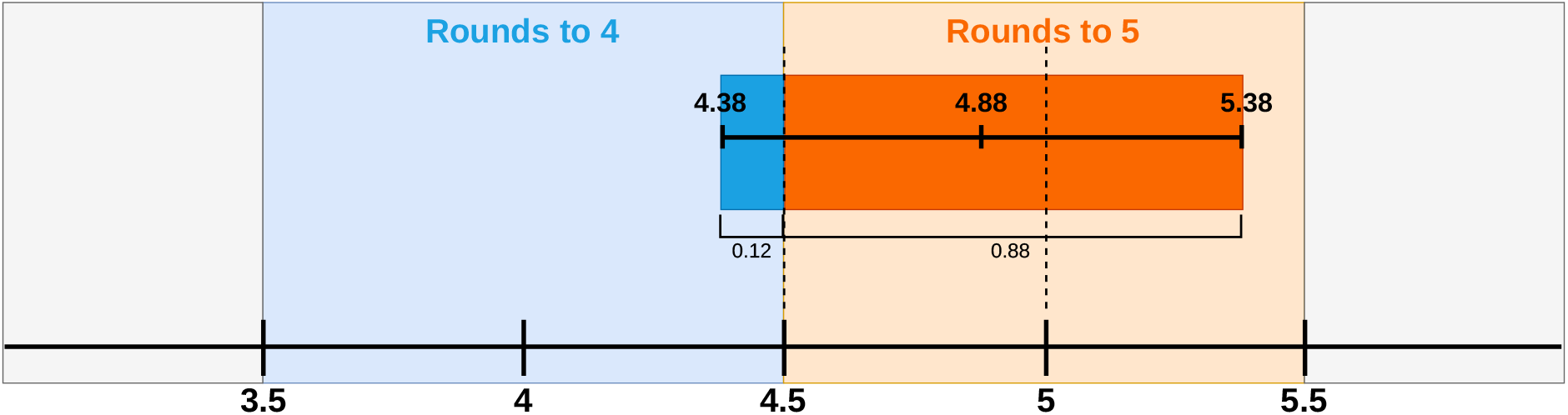
Number-line diagram illustrating why single-step ADU transitions produce stream deltas of either ⌊*α*⌋ or ⌈*α*⌉, and in what proportion. When an ADU value changes by 1 between subsequent samples, the unrounded delta in stream units (Δ*S*) would be exactly 1 · *α* = 4.88. However, the actual stream unit delta is round(*α* + *ϵ*_*n*_), where *ϵ*_*n*_ ∈ [−0.5, 0.5] is the residual rounding carried in from the previous sample. The actual quantity being rounded is therefore *α* ± *ϵ*_*n*_, which can land anywhere in the window [*α* − 0.5, *α* + 0.5] depending on the value of *ϵ*_*n*_. For *α* = 4.88, the window [*α* − 0.5, *α* + 0.5] = [4.38, 5.38] overlaps the “rounds to 4” zone on a small range of values from 4.38 to 4.5, which has width = 0.12. When *α* + *ϵ*_*n*_ falls in this sub-interval, the delta rounds to 4. The window overlaps the “rounds to 5” zone on a larger range, from 4.5 to 5.38, which has width = 0.88. When *α* + *ϵ*_*n*_ falls here, the delta rounds to 5. If *ϵ* is approximately uniformly distributed, then the width of the windows which round to either 4 or 5 represent the probabilities of a change in 1 ADU mapping to 4 (0.12) or 5 (0.88). The probability split is determined by where *α* sits within the unit interval, and for large *N, α* is approximated by the mean of observed stream unit deltas for all single ADU steps.

This is graphically illustrated for a *k* = 1 unit ADU step in Figure 4. As a result, and as shown in Figure 2, for *α* = 4.88, Δ*S* can only take on values of {0, ±4, ±5, ±9, ±10, ±14, ±15, …}, which correspond to the integers bracketing *kα* for each integer ADU step *k*. The values {±1, ±2, ±3, ±6, ±7, ±8, ±11, ±12, ±13, …} are never observed. This pattern with gaps in signal deltas is the signature of double quantization [13, 14]. It also confirms that the encoder from ADUs to stream units did not introduce any significant filtering, dithering, or smoothing, and therefore, all information needed to invert the encoding back to ADU values is preserved in the stream values. Figure 2 also illustrates that the augmented limb leads, aVR, aVL, and aVF, which exist in half-ADU space since they involve division by 2 (see Eq. 15 below), are rounded by MUSE prior to plotting. If any leads existed as non-integer ADU values at the time of conversion to stream units, the observed values for Δ*S* would be different (see Section 2.13.1 and Supplemental Section S.4).

#### 2.10.3. Formulation of the ADU Reconstruction Problem

We start by looking at the un-rounded conversion (*Y*_*n*_) of ADUs into stream units based on the stream-to-ADU relationship (*α*) and the offset introduced during PDF generation to position the waveform at a specific page location (*B*):

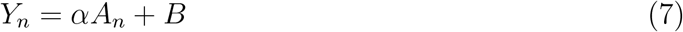

The stream units (*S*_*n*_) that are encoded in the PDF must be integers, and are therefore the rounded values of *Y*_*n*_:

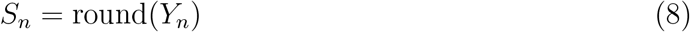

We define the rounding error between *S*_*n*_ and its true value *Y*_*n*_ as:

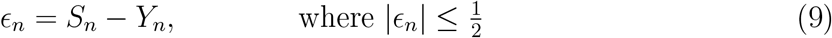

The encoding relationship used by the PDF generator is therefore:

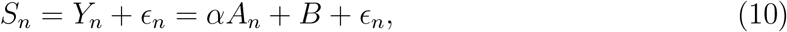

Our goal is to reconstruct *A*_*n*_ from *S*_*n*_, *α*, and *B*. To do that, however, we need to determine the effect of *ϵ*_*n*_.

Inverting stream units to ADUs is actually relatively straightforward if *α* and *B* are known. In that case, we subtract *B*, divide by *α*, and round each side of Eq. 10:

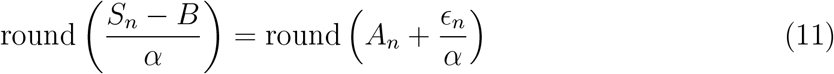

Because 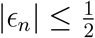, 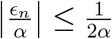, and for *α >* 1 (which we will subsequently show is a requirement of any bit-exact reconstruction), 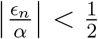, and the rounding operation therefore removes the 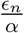 term. As *A*_*n*_ is an integer, it is not affected by the round operation.

As a result, as long as *α >* 1 we can *exactly* reconstruct *A*_*n*_:

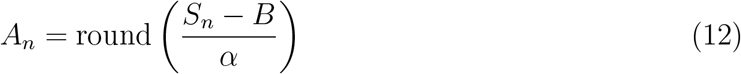

To obtain physical units (*V*), we multiply by the mV per ADU relationship *m*:

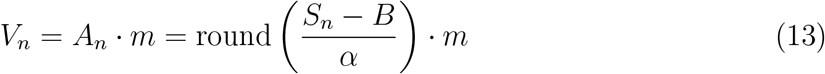

Note how Eq. 13 differs from Eq. 2 due to the round operation prior to multiplication by *m*, even though the variables used in each equation (*α* = *m* · *g*_*v*_ · *G*, Eq. 4) are the same.

### 2.11. Calculating ECG Sampling Frequency

ECG sampling frequency (*f*_*s*_) is calculated based on the number of samples in the signal (*N*) and the total duration of the signal:

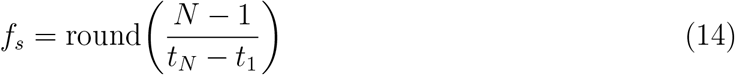

where *t*_*N*_ − *t*_1_ is the duration of the signal in seconds obtained from Eq. 1. The software also checks that the inter-sample time interval is uniform to within one stream unit (1/2500 s ≈ 0.4 ms for 25 mm/s), which is the quantization floor imposed by the integer-valued stream coordinates. The first sample of each lead is shifted to time *t* = 0. Note that any quantization of the horizontal axis is purely a display artifact and not a resampling of data. As a result, sample *n* in the PDF and sample *n* in the XML correspond to the same underlying recorded sample even if the absolute times of sample *n* differ very slightly between XML and PDF.

### 2.12. Clipping Detection

One of the major causes of disagreement between the PDF and ground truth XML waveforms is secondary to “clipping” of the PDF waveform. Clipping occurs when stream coordinates exceed the minimum or maximum that can be displayed on the PDF page. There is no method that can recover the clipped signals, as the clipped information is not included in the PDF. By evaluating the minimum and maximum stream values in over 376,000 leads, we empirically determined that stream values that would otherwise fall below 450 stream units or above 21,150 stream units are clipped by the MUSE PDF renderer to those limit values. An example of the effect of clipping is shown in Figure 5. The software identifies any lead in which any stream unit equals 21,150 or 450 and flags that ECG lead as having clipping which might significantly reduce waveform reconstruction accuracy. Sample-level clip masks, number of clipped samples per lead, and the total number of clipped samples over the full ECG are returned alongside the digitized signals so that downstream analyses can exclude or annotate affected leads.

**Figure 5:**
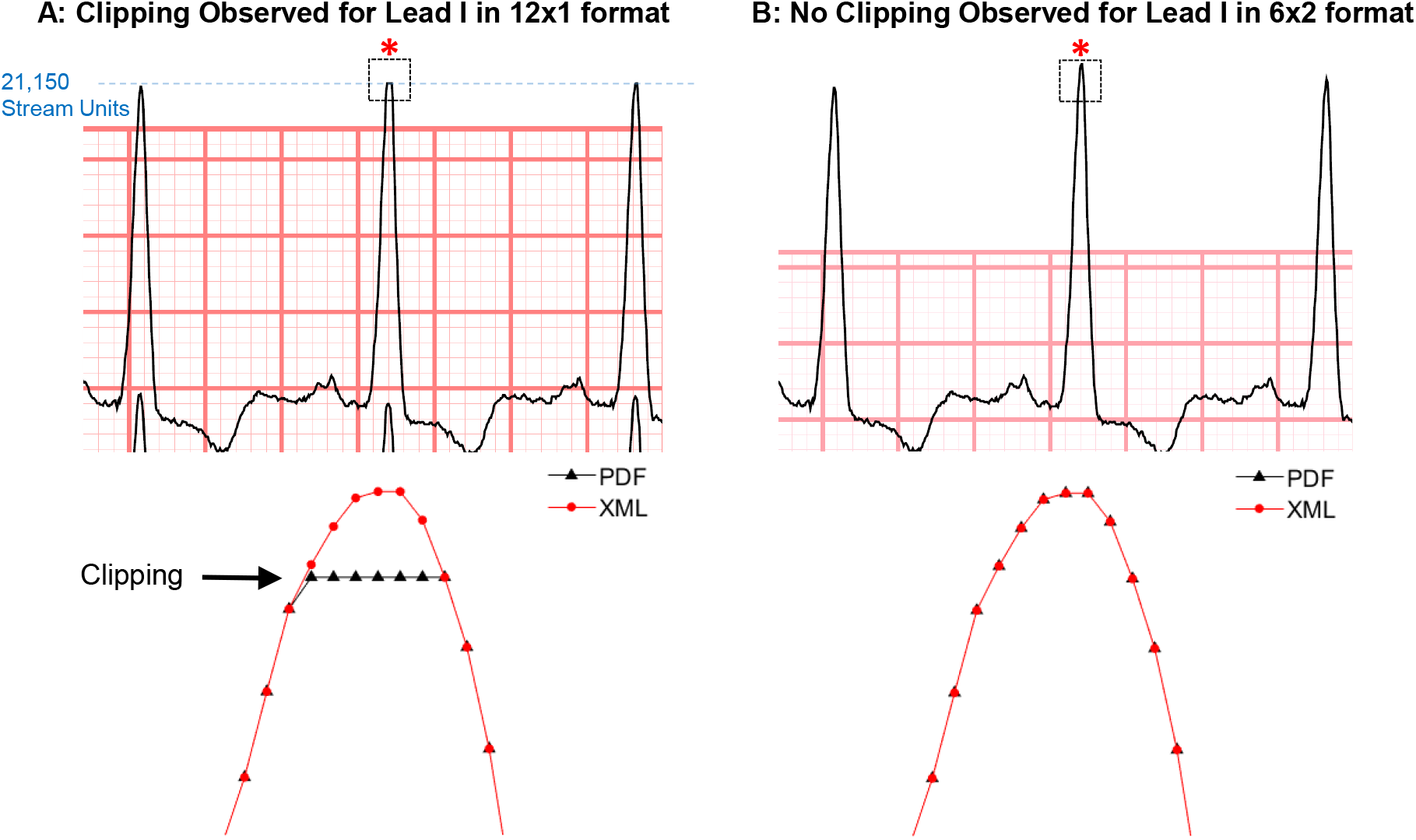
Illustration of how clipping results in loss of ECG waveform data. **Panel A** shows lead I in a 12 ×1 format ECG where the R wave peak is clipped in some beats. This occurs because the waveform stream value exceeds 21,150, which is the maximum value that can be displayed. The square area is zoomed in and shows that once the stream units reach 21,150, the PDF recovered waveform (black) is clipped at this value resulting in significant error when compared to the waveform obtained from the XML (red). **Panel B** shows how clipping depends on the PDF ECG display format, as the same ECG data displayed in 6 × 2 layout does not have any clipping and the red (XML) and black (PDF) signals are identical.

### 2.13. Augmented Lead Reconstruction

MUSE XML files contain the 8 independent leads I, II, and V1-V6. Leads III, aVR, aVL, and aVF must be derived from leads I and II using Einthoven’s and Goldberger’s equations:

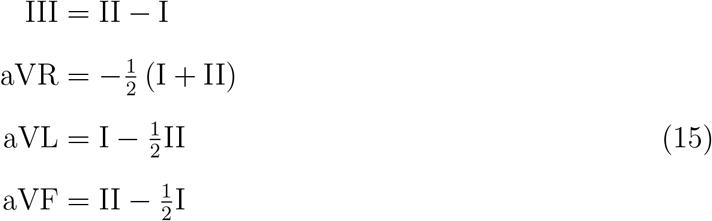

In contrast, the PDF contains data on all 12 leads. The software has 2 options for extracting the derivable leads III, aVR, aVL, and aVF. In “pdfleads” mode, the 4 derivable leads are extracted directly from the encoded PDF values. In “calcleads” mode, the 4 derivable leads are calculated from leads I and II in the same way as is done when processing the equivalent XML file using Eq. 15. When the derivable leads are calculated, any clipped samples in leads I or II are propagated to all leads derived from them; although these leads do not necessarily have stream units outside the min/max values, the accuracy of their calculated stream values will likewise be affected if lead I or II is clipped.

#### 2.13.1. Definition of Ground Truth for the Augmented ECG Leads

As noted, MUSE exported raw waveforms in XML format do not encode data for leads III, aVR, aVL, and aVF. The ground truth for these leads, therefore, must be derived from XML leads I and II. Lead III (III = II - I) ADU values are always integers, but leads aVR, aVL, and aVF exist in half-ADU values (see Eq. 15 above).

We know from the analysis of stream deltas (see Section 2.10.2 and Figure 2) that only integer ADU values are encoded into stream units, and prior to converting aVR, aVL, and aVF into stream units for PDF display, MUSE uses round-half-toward-zero on the half-ADU values of aVR, aVL, and aVF. There are therefore 2 possible “ground truth” values for aVR, aVL, and aVF; either the calculated values obtained directly from Eq. 15 that exist in half ADU values, or MUSE’s pre-encoding rounded ADU integer, which differs from the unrounded values by ≤ 0.5 ADU. For layouts where aVR, aVL, and aVF are simultaneous with leads I and II (all layouts except 3 ×4), exact agreement with either augmented lead ground truth definition is achievable. For 3×4 layouts, however, aVR, aVL, and aVF (displayed between 2.5-5 sec) are not simultaneous with leads I and II (displayed between 0-2.5 sec), and as a result, “calcleads” mode cannot reconstruct aVR, aVL, and aVF at the time they are actually displayed. As a result, to accurately reconstruct aVR, aVL, and aVF in 3×4 formats, “pdfleads” mode should always be used. Therefore “calcleads” mode assumes the half-ADU values for aVR, aVL, and aVF as ground truth, while “pdfleads” assumes the round-towards-zero values as ground truth, with neither being “correct”. Further information is available in the Supplement Section S.4.

### 2.14. Rhythm Strip Matching

In PDF formats such as 3×4 2.5-second leads with 1 or 3 10-second rhythm strips at the bottom of the page, the rhythm strip leads displayed and their order are not fixed. Each rhythm strip polyline is identified by matching it to its label in the data stream. Any lead displayed as a rhythm strip can be extracted, and the software does not require any specific rhythm strip ordering.

### 2.15. Certification of Waveform Reconstruction

Waveform reconstruction is bit-exact when the values of *α* and *B* used in Eq. 12 are the same values as which were used by MUSE to encode the PDF. The program uses 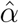 which is determined from the PDF geometry and the assumed MUSE voltage sensitivity (see Section 2.9 and Eq. 4), and 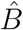 which is determined from the calibration signal baselines (see Section 2.7). Because the voltage-to-ADU relationship (*m* in mv/ADU, which is used to calculate the stream-to-ADU relationship, *α*) is not explicitly encoded in the PDF, and the baseline offsets are likewise not explicitly defined in the PDF datastream, an incorrect value of either 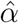 or 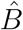 could result in a silent reconstruction error when the corresponding XML signal is unavailable to validate that a waveform reconstruction was correct.

We therefore developed three complementary certification tests that determine whether the values of 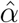 and 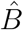 used by the software for waveform reconstruction are consistent with the observed stream values generated using the true values, *α* and *B*, which we cannot directly obtain. All three tests utilize exact integer arithmetic and therefore do not require any numerical tolerance. When all three tests pass, they provide a ground truth-free certificate of bit-exact reconstruction under the stated MUSE encoding model (Eq. 10). Full derivations are provided in the Supplement Section S.5, with brief descriptions provided here.

#### 2.15.1. Residue Width Test, W

The residue width test is used to assess if the value of 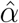 used by the program is consistent with the value of *α* which was used to encode the PDF. For this section we will assume that the value of the offset is correct and therefore 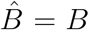. Under the encoding model (Eq. 10) we have:

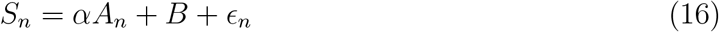

We do not know *α*, and need to test if our value of 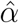 is consistent with the value of *α*. To do this, we define 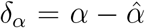, substitute 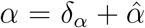 into Eq. 16, and then take modulo 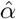 of both sides of the equation:

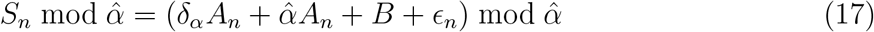

This has the effect of wrapping the stream units around a circle of circumference 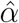 as shown in Supplemental Figure 2. Taking mod 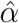 of both sides of the equation makes the 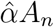 term vanish as it is an integer multiple of 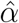 and therefore reduces to 0 mod 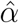. As a result:

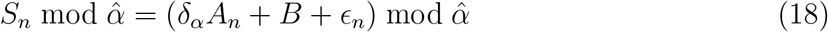

If 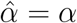, *δ*_*α*_ = 0, and the remaining *δ*_*α*_*A*_*n*_ term also vanishes leaving:

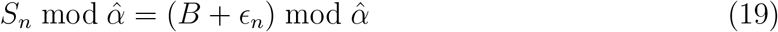

*B* is a constant, and 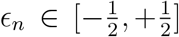, so the residues on the right side of the equation collapse onto a circular arc centered on *B* mod 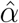 with a maximum width of 1 stream unit due to the range of *ϵ*_*n*_ (see Supplemental Figure 2). We define *W* as the length of the shortest arc that contains all of the residues, and based on the bound of *ϵ*_*n*_ it can be seen that in the case where 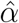 is consistent with *α, W* ≤ 1 stream unit (see Figure 6A). Because 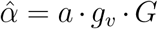 is a product of the MUSE mV-to-ADU relationship (*m*) with two integers read from the PDF (*g*_*v*_ and *G*; see Eq. 4), it is therefore a rational number which can be written in lowest terms as 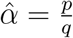 for non-zero integers *p* and *q*. As a result, the residues are also quantized (in steps of 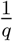), and due to this quantization, the bound tightens to *W* ≤ (*q* − 1)*/q <* 1 stream units. This bound can be further refined to avoid any floating point errors that might push the test to incorrectly pass or fail at the boundary (see Supplement Section S.5 for further details of this derivation).

**Figure 6:**
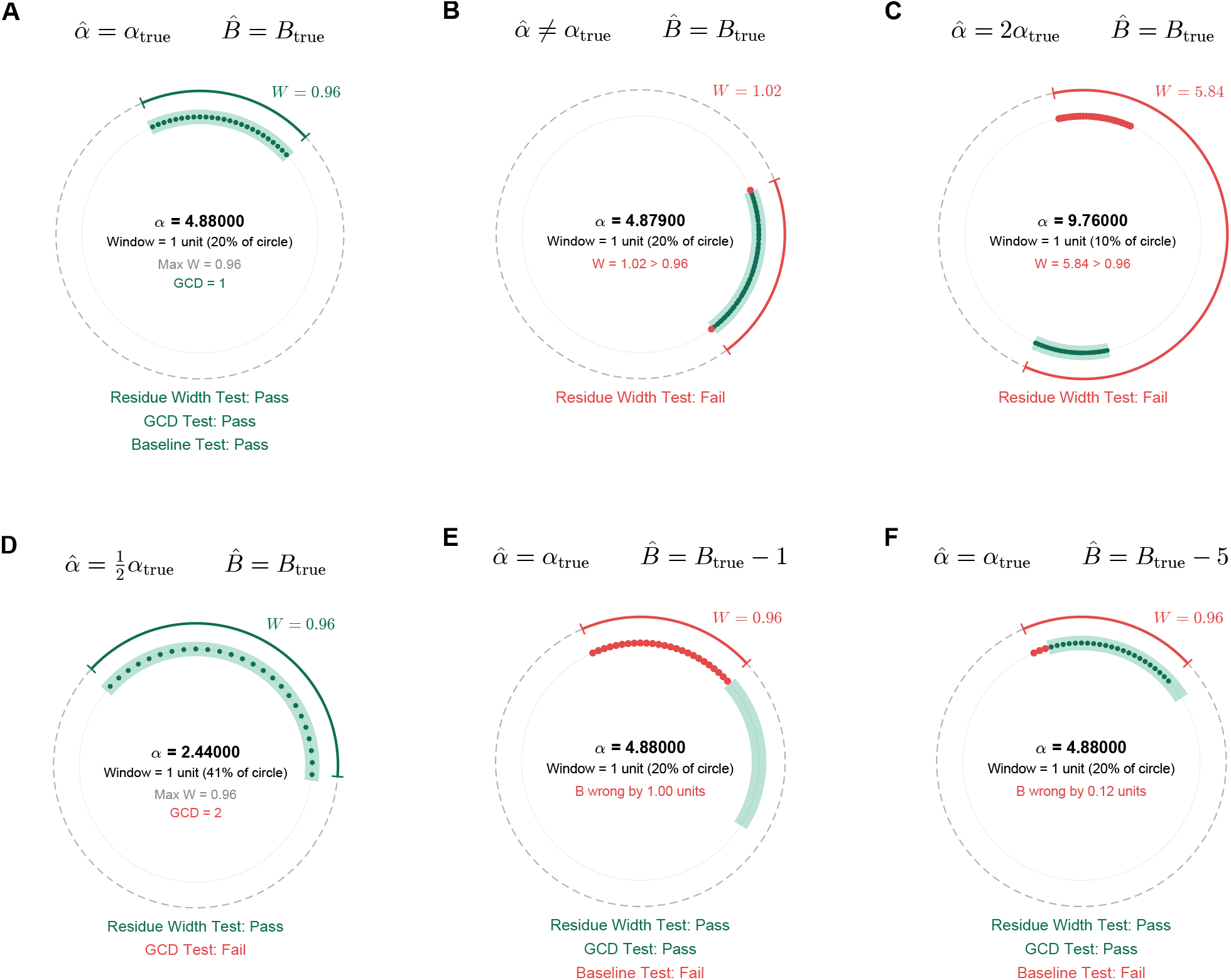
Examples of certification tests results (see Section 2.15 and Supplement Section S.5 for further details). In each panel the light green shaded area is the window where residues are expected to exist. Actual quantized residues are represented by green dots when they are located within the expected window, and red dots when when located outside of the expected window. The width of the window is noted by *W* which should be ≤0.96 to pass the window residue test for all MUSE encoded PDFs. **Panel A**: When 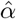 and 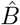 are the same as the values used for PDF encoding, the residues all exist within the expected range, with width 0.96. This passes the residue width, GCD, and baseline tests. **Panel B**: If 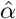 is incorrect (in this example even very slightly), some of the residues fall outside of the expected window, and *W >* 0.96. This fails the residue width test. **Panel C**: When 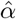 is an integer multiple *k* of *α*_true_, the residues occupy *k* regions, equidistributed around the circle. In this case, as *k* = 2, there are 2 distinct areas where residues cluster. Only 1 of the areas is the predicted area, and as a result *W >* 0.96. This fails the residue width test. **Panel D**: When 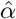 is an integer submultiple of *α*_true_, this discrepancy cannot be detected by the residue width test. As can be seen, the residues occupy a larger percent of the circle circumference, but *W* = 0.96. This passes the residue width test, but fails the GCD test (see Figure 7) so the lead is not incorrectly certified. **Panel E**: When 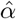is same as the value used for PDF encoding, but 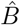 is incorrect, the residues occupy the correct width (*W* = 0.96), but the residues are located in the wrong sector of the circumference. In this case, since 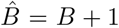, they are shifted 1 full stream unit around the circle from where they are expected. This passes the residue width and GCD tests, but fails the baseline test. **Panel F**: Another example where 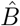 is incorrect. In this case even if the shift rotates around the circle, it will not line up perfectly with the expected residue location, and there will always be some residues that are outside the predicted area, leading to failure of the baseline test.

If 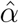 is *not* consistent with *α*, the *δ*_*α*_*A*_*n*_ term in Eq. 18 does *not* vanish, and unlike as shown in Eq. 19 when *δ*_*α*_ = 0, the residues may no longer be confined to the expected ≤ 1 stream unit arc centered on *B* mod 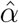. Instead, the residues are now additionally displaced based on the value of *δ*_*α*_*A*_*n*_ mod 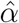. As a result, as *A*_*n*_ changes, the additional term in Eq. 18 can increase the width of the residues beyond that expected when *δ*_*α*_ = 0, leading to *W >* (*q* − 1)*/q* as shown in Figure 6B-C and Supplemental Figure 2. Therefore, observing *W >* (*q* − 1)*/q* indicates that 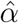 is *not* consistent with *α*, while observing *W* ≤ (*q* − 1)*/q* indicates that 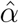 passes the residue width test. Importantly, passing this test with *W* ≤ (*q* − 1)*/q* does not always prove that 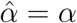, and additional testing, as discussed below, is required. The residue width test is highly sensitive to mismatch between 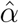 and *α*, although failure does not always indicate bit-exact reconstruction will also fail, as the reconstruction errors may not be large enough to cross a rounding boundary (see Supplemental Section S.6). Further details on the residue width test derivation are available in Supplement Section S.5.

#### 2.15.2. Greatest Common Divisor (GCD) Test

Although the residue width test is very sensitive to errors in 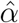, it is unable to detect if 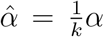 for integer *k*. This specific error reconstructs every sample as *kA*_*n*_ but leaves the residue structure unchanged because, in this case, 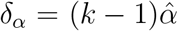, and substituting into Eq. 18:

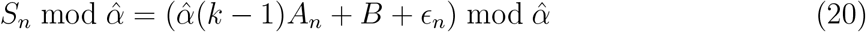

Because *A*_*n*_ and *k* − 1 are integers, the 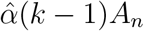 term vanishes mod 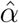 leaving

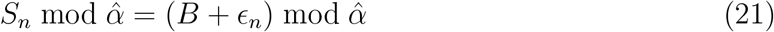

This is the same as Eq. 19 despite 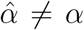. Therefore, if 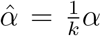 for integer *k* ≥ 2 it will pass the residue width test despite this inconsistency. We therefore need a second test for this specific scenario. To detect if 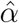 is an integer submultiple of *α*, we assess the greatest common divisor (GCD) of the non-zero reconstructed differences *Â*_*n*+1_ − *Â*_*n*_. The integer submultiple error scales every sample by *k*, so *Â*_*n*_ = *kA*_*n*_, and

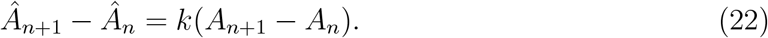

as a result, *all* non-zero differences are divisible by *k*. If the GCD of *Â*_*n*+1_ − *Â*_*n*_ equals 1, an integer submultiple error in 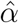 is excluded and the GCD test is passed. This is illustrated in Figures 6D and 7, with and additional information available in Supplement Section S.5.

#### 2.15.3. Baseline Test

Given a certified value of 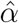 which passes the residue width and GCD tests, as noted in Eq. 19, the residues must be confined to a ≤ 1 stream unit arc, with the center of the residue arc determined by the extracted offset 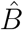 which may not equal the true offset *B*. An error in the assumed baseline 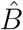 therefore displaces the location of the expected residue window, and the actual calculated residues, which are calculated from *S*_*n*_ mod 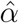, with *S*_*n*_ encoded with the correct offset *B*, will not fall within the circular arc defined by 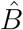 mod 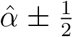. If *any* calculated residue falls outside of the window determined by 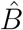 the test fails as shown in Figure 6E-F. The baseline test is evaluated only when the prior two tests on 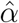 pass because the predicted window is undefined otherwise. See Supplement Section S.5 for additional information.

#### 2.15.4. Waveform Certification

A reconstructed waveform is certified bit-exact when all three certification tests pass. Certification establishes that the observed stream values are consistent with a PDF encoder (Eq. 10) which used the extracted stream-to-ADU sensitivity 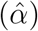 and baseline 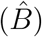, and that under those parameters the ADU recovery is therefore bit-exact. Failure of a test does not by itself imply that reconstruction is not bit-exact, but indicates that exactness cannot be guaranteed from the PDF stream unit structure alone. The residue width test, in particular, is far more sensitive to 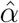 error than bit-exactness requires (see Supplement Section S.6), and a sufficiently small error in 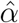 may not move any sample across an ADU rounding boundary. Further discussion about how much 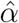 must deviate from *α* and how much 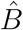 must deviate from *B* to violate bit-exact reconstruction are available in Supplement Sections S.6 and S.7, respectively.

### 2.16. Data Output

The software returns 2 structures: `signals` contains the 12 standard ECG leads in units of mV or ADUs, any matching rhythm strips, and clipping totals and results of certification for easy identification of possible reconstruction issues, and `info` which contains additional information about PDF processing, clipping, and certification. Further details of what is contained in the output structures can be found in Table 6. Recovered waveforms can optionally be output as a .csv file as well.

### 2.17. Assessment of Waveform Reconstruction Accuracy and Certification

To assess the accuracy of ECG waveform reconstruction, all 12 lead waveforms reconstructed from the PDF were compared to the same 12 lead waveform data obtained from its matched XML file. As leads with clipping have loss of data that can never be recovered, leads with clipping detected were excluded from the main analysis with results reported separately. All leads were included regardless of noise or artifact. Both definitions of aVR, aVL, and aVF ground truth using “calcleads” mode and “pdfleads” mode, were tested separately. We tested 5 different PDF layout formats and 3 gains (5, 10, and 20 mm/mV).

The primary method of assessing waveform recovery accuracy was determining if the integer ADU values reconstructed from the PDF were identical (bit-exact) to the matched XML integer ADU values. Although ECG analysis uses physical units of mV, because there is a linear relationship between ADUs and mV (1 ADU = 4.88 *μ*V), confirming identical ADUs also confirms that the physical units recovered from the PDF are identical to the XML-derived physical units. However, because ADUs are integers, comparing ADUs rather than mV avoids issues related to floating-point precision. For PDF layout formats where a full 10-seconds of every lead waveform was not printed on the PDF (all formats except 12×1), only the visible intervals on the PDF were compared to the equivalent intervals in the XML. In 3 ×4 format with 1 or 3 rhythm strips, the rhythm strip lead was identified, and the full 10-seconds of rhythm strip were compared to the full 10-seconds of equivalent XML data. As the accuracy of waveform reconstruction is independent of patient, multiple ECGs per patient were allowed.

Secondary analyses included assessing the accuracy of direct stream units-to-mV conversion (Eq. 2) and the effect of clipping by calculating the maximum error, MAE, and RMSE for each of the 12 leads with the PDF and XML signals aligned at time = 0. We assessed the performance of the three bit-exact certification tests in response to varying values of 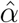 and 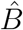, to determine how errors in these values affected waveform reconstruction accuracy, and if leads with these incorrect 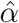and 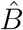 correctly failed certification. Statistical analysis was performed in MATLAB R2022a.

## 3. Results

### 3.1. Study Population

The study included a total of 1,632 ECGs with matched PDF and XML output from MUSE v10, and 2,336 ECGs with matched PDF and XML output from MUSE v9. The MUSE v10 PDFs were available in 5 formats and 3 gains (24,480 total PDFs and 313,344 total leads), and were therefore used as the primary dataset, while the MUSE v9 PDFs were only available in 12 ×1 and 3 ×4 +3 formats, both at 10 mm/mV (4,672 total PDFs and 63,072 total leads). In total, 29,152 PDFs and 376,416 leads were processed. Detailed demographics and medical history were not available, but in the primary cohort, mean age was 65 ± 17 years, and 50% were male. Age and sex were not available for the MUSE v9 ECGs which had been previously deidentified. In total, 29,152 PDFs were used to evaluate the reconstruction algorithm, and the distribution of ECGs per format and gain is shown in Table 1 and Supplemental Table 2.

**Table 1:** Summary of bit-exact waveform recovery for different PDF layouts and voltage gains in 1,632 ECGs. Numbers represent number of leads with bit-exact reconstruction, the total number of leads processed (after excluding clipped leads), and the percent of the total. After excluding clipping, bit-exact recovery was present for 100% of leads in all formats and gains. *calcleads format does not reconstruct leads aVR, aVL, and aVF in any 3×4 format because these leads are not simultaneous with leads I and II, and as a result there are fewer total leads assessed for 3×4 formats in calcleads mode.

| Layout | Mode | Gain |  |  |
| --- | --- | --- | --- | --- |
|  |  | 5 mm/mV | 10 mm/mV | 20 mm/mV |
| $12 \times 1$ | calcleads* | 19560/19560<br>100% | 19474/19474<br>100% | 17175/17175<br>100% |
|  | pdfleads | 19568/19568<br>100% | 19541/19541<br>100% | 18899/18899<br>100% |
| $6 \times 2$ | calcleads* | 19578/19578<br>100% | 19577/19577<br>100% | 19517/19517<br>100% |
|  | pdfleads | 19578/19578<br>100% | 19571/19571<br>100% | 19431/19431<br>100% |
| $3 \times 4 + 3$ | calcleads* | 19579/19579<br>100% | 19537/19537<br>100% | 19421/19421<br>100% |
|  | pdfleads | 24475/24475<br>100% | 24433/24433<br>100% | 24316/24316<br>100% |
| $3 \times 4 + 1$ | calcleads* | 16318/16318<br>100% | 16317/16317<br>100% | 16269/16269<br>100% |
|  | pdfleads | 21214/21214<br>100% | 21213/21213<br>100% | 21163/21163<br>100% |
| $3 \times 4 + 0$ | calcleads* | 14686/14686<br>100% | 14669/14669<br>100% | 14484/14484<br>100% |
|  | pdfleads | 19582/19582<br>100% | 19564/19564<br>100% | 19324/19324<br>100% |

**Table 2:** Errors associated with direct stream to mV conversion. The maximum error corresponds to 0.5 stream unit. Abbreviations: MAE = mean absolute error, RMSE = root mean squared error.

| **Gain**<br>(mm/mV) | **MAE**<br>(μV) | **RMSE**<br>(μV) | **Max error**<br>(μV) |
| --- | --- | --- | --- |
| 5 | $0.496 \pm 0.006$ | $0.575 \pm 0.005$ | 0.96 |
| 10 | $0.248 \pm 0.007$ | $0.288 \pm 0.007$ | 0.48 |
| 20 | $0.124 \pm 0.001$ | $0.145 \pm 0.001$ | 0.24 |

### 3.2. Digitization Accuracy Results

All 29,152 PDF files (100%) were successfully processed by the software without errors. Results comparing PDF reconstructed waveforms to the raw XML waveforms are shown in Table 1 and Supplemental Table 2. After excluding leads with clipping (see Section 3.4), 100% of reconstructed waveforms, using both definitions of ground truth for the augmented leads, were bit-exact to the equivalent raw ADU data contained in XML files. Detailed information on each lead in each ECG format and gain are shown in Supplemental Tables 3 and 4.

### 3.3. Direct Stream Units to mV Conversion

Table 2 shows residual errors in *μ*V averaged over all leads when directly converting stream units to voltage for 12×1 format in “pdfleads” mode at gains of 5, 10, and 20 mm/mV. As expected, 0% of leads had complete and exact agreement with the XML-derived voltage values. Errors were inversely related to gain and correlated with the expected error for a quantized error spanning ±0.5 stream unit, which has a different conversion factor to voltage units at each gain. Results were consistent with prior reports [9]. The observed values of MAE and RMSE suggest that the residual errors were entirely due to rounding. Further details are available in Supplement Section S.8.

### 3.4. Waveform Clipping Results

The overall frequency of ECGs with at least 1 clipped lead with different PDF layouts and gains are shown in Table 3. The prevalence of clipping varied by ECG format and gain. Gains of 5 mm/mV had the lowest rates of clipping (*<* 0.4% overall), while gains of 20 mm/mV had the highest rates (2.9 − 33.0% in different formats). The 12 × 1 format had the highest rates of clipping when compared to the other formats, which is expected given the 12×1 format has the least space for displaying ECG waveforms away from the extremes of the page. Notably, all ECGs with clipping did not necessarily have loss of *physiological* information, as the clipped samples were often noise/artifact or large pacing spikes, although there is no way to know this without inspection of the ECGs that were flagged. Detailed data on the prevalence of clipping in specific leads within specific PDF formats and gains are provided in Supplemental Tables 5 and 6, which overall show that leads near the extremes of the page boundaries for the specific format layout were most likely to clip. In “calcleads” mode the augmented leads have different rates of clipping when compared to “pdfleads” mode even though clipping is determined based on the printed PDF, because the augmented leads could inherit the increased rate of clipping from lead I, or be “rescued” if an augmented lead clipped but then was recalculated without clipping from leads I and II.

**Table 3:** Number of ECGs with clipping in any lead. Numbers represent number of ECGs with clipping, total number of ECGs, and percent of the total. As clipping is a feature of the PDF, reported values are only for pdfleads mode.

| PDF Layout | Gain |  |  |
| --- | --- | --- | --- |
|  | 5 mm/mV | 10 mm/mV | 20 mm/mV |
| $12 \times 1$ | 6/1632<br>0.37% | 28/1632<br>1.72% | 538/1632<br>32.97% |
| $6 \times 2$ | 2/1632<br>0.12% | 8/1632<br>0.49% | 115/1632<br>7.05% |
| $3 \times 4 + 3$ | 2/1632<br>0.12% | 44/1632<br>2.70% | 147/1632<br>9.01% |
| $3 \times 4 + 1$ | 1/1632<br>0.06% | 2/1632<br>0.12% | 47/1632<br>2.88% |
| $3 \times 4 + 0$ | 1/1632<br>0.06% | 19/1632<br>1.16% | 222/1632<br>13.60% |

Clipping had a profound effect on the error of waveform recovery as shown in Supplemental Table 7 which shows errors associated with clipping in the 12 ×1 format at a gain of 10 mm/mV. Leads with clipping were, by definition, not reconstructed bit-exactly since clipping destroys information beyond the clipping boundary, but the magnitude of errors were widely variable, depending on both the magnitude of clipping and how many samples actually clipped. Some of the extreme errors were due to clipping of non-physiological noise/artifact.

### 3.5. Certification of Bit-Exact Reconstruction

As shown in Table 4, 100% of ECG leads in all formats and gains without clipping in the MUSE v10 cohort were reconstructed bit-exactly and passed all 3 certification tests. In the MUSE v9 cohort all leads without clipping were also reconstructed bit-exactly, but 2 ECG leads from 2 separate ECGs had an indeterminate certification despite being bit-exact reconstructions. Review of these 2 leads demonstrated that the cause of indeterminate certification was that they were disconnected leads with no data. This indeterminate classification is appropriate, as the disconnected leads have no range, the residues therefore occupy a single point and the lead is appropriately flagged.

**Table 4:** Certification test results by PDF layout, pooled over gains of 5, 10, and 20 mm/mV. Numbers represent number, total, and percent of the total. ECG-level counts include records with no clipping on any lead, and lead-level counts include leads with no clipping, so an ECG with 1 clipped lead still contributes its remaining leads.

| Layout | Mode | ECGs |  |  | Leads |  |  |
| --- | --- | --- | --- | --- | --- | --- | --- |
|  |  | Certified | Failed | Indeterminate | Certified | Failed | Indeterminate |
| $12 \times 1$ | calcleads | 4,329/4,329<br>100% | 0/4,329<br>0% | 0/4,329<br>0% | 56,280/56,280<br>100% | 0/56,280<br>0% | 0/56,280<br>0% |
|  | pdfleads | 4,329/4,329<br>100% | 0/4,329<br>0% | 0/4,329<br>0% | 58,079/58,079<br>100% | 0/58,079<br>0% | 0/58,079<br>0% |
| $6 \times 2$ | calcleads | 4,843/4,843<br>100% | 0/4,843<br>0% | 0/4,843<br>0% | 58,672/58,672<br>100% | 0/58,672<br>0% | 0/58,672<br>0% |
|  | pdfleads | 4,771/4,771<br>100% | 0/4,771<br>0% | 0/4,771<br>0% | 58,580/58,580<br>100% | 0/58,580<br>0% | 0/58,580<br>0% |
| $3 \times 4 + 3$ | calcleads | 4,708/4,708<br>100% | 0/4,708<br>0% | 0/4,708<br>0% | 58,608/58,608<br>100% | 0/58,608<br>0% | 0/58,608<br>0% |
|  | pdfleads | 4,708/4,708<br>100% | 0/4,708<br>0% | 0/4,708<br>0% | 73,313/73,313<br>100% | 0/73,313<br>0% | 0/73,313<br>0% |
| $3 \times 4 + 1$ | calcleads | 4,846/4,846<br>100% | 0/4,846<br>0% | 0/4,846<br>0% | 48,904/48,904<br>100% | 0/48,904<br>0% | 0/48,904<br>0% |
|  | pdfleads | 4,846/4,846<br>100% | 0/4,846<br>0% | 0/4,846<br>0% | 63,590/63,590<br>100% | 0/63,590<br>0% | 0/63,590<br>0% |
| $3 \times 4 + 0$ | calcleads | 4,684/4,684<br>100% | 0/4,684<br>0% | 0/4,684<br>0% | 43,839/43,839<br>100% | 0/43,839<br>0% | 0/43,839<br>0% |
|  | pdfleads | 4,654/4,654<br>100% | 0/4,654<br>0% | 0/4,654<br>0% | 58,470/58,470<br>100% | 0/58,470<br>0% | 0/58,470<br>0% |

### 3.6. Sensitivity of Certification Tests

Table 5 shows results from sensitivity analyses of the certification tests using different values of 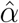and 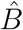 instead of the true encoding values on 1,632 XML/PDF pairs in 12×1 format at 10 mm/mV in “pdfleads” mode. Certification failed, as expected, in 100% of waveforms when *α* = 4.88 but reconstruction used 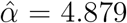 (100% failure of the residue width test), 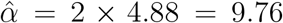 (100% failure of the residue width test), or 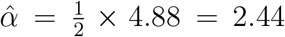 (passes the residue width test but then has 100% failure of the GCD test). Shifting 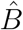 by 1 stream unit resulted in 100% failure of the baseline test, although bit-exact reconstruction still occurred (see Supplement Section S.7). When 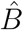 was shifted by ≥ 2 stream units, there was 100% failure of the baseline test and failure of bit-exact reconstruction (see Figure 6 and Supplemental Figures 7 and 8).

**Table 5:** Summary of perturbation results for 1,632 ECGs in 12 × 1 format at a gain of 10 mm/mV. At baseline (row 1), where 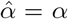 and 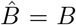, 100% of ECG waveforms pass all 3 certification tests and 100% of all waveforms are reconstructed bit-exact. Errors in 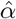 or 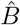 result in failure of certification, although failure of certification does not always result in failure of bit-exact reconstruction. For example, at gain 10 mm/mV, a 1 stream unit error in 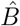(row 6) results in 0% certification, but 100% of waveforms are still reconstructed bit-exact, while larger errors in 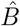 (rows 7-8) fail certification and bit-exact reconstruction. A very small error in 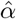 (row 2) results in certification failure of 99.8% of waveforms even though 100% of waveforms are reconstructed bit-exact, while larger errors in 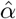 result in higher certification failure rates and lower rates of bit-exact reconstruction (rows 3-5).

| Condition | W range | Pass W | Pass GCD | Pass B | Certified | Bit-exact |
| --- | --- | --- | --- | --- | --- | --- |
| Baseline | 0.96 | 19,541/19,541<br>100% | 19,541/19,541<br>100% | 19,541/19,541<br>100% | 19,541/19,541<br>100% | 19,541/19,541<br>100% |
| $\hat{\alpha} = 4.8793$ | 0.9656-2.386 | 386/19,541<br>2.0% | 19,541/19,541<br>100% | 45/386<br>11.7% | 45/19,541<br>0.2% | 19,541/19,541<br>100% |
| $\hat{\alpha} = 4.87$ | 1.08-4.86 | 0/19,541<br>0% | 19,541/19,541<br>100% | N/A | 0/19,541<br>0% | 13,009/19,541<br>66.6% |
| $\hat{\alpha} = \frac{1}{2} \times 4.88$ | 0.96 | 19,541/19,541<br>100% | 0/19,541<br>0% | N/A | 0/19,541<br>0% | 0/19,541<br>0% |
| $\hat{\alpha} = 2 \times 4.88$ | 5.72-5.84 | 0/19,541<br>0% | 19,541/19,541<br>100% | N/A | 0/19,541<br>0% | 0/19,541<br>0% |
| $\hat{B} = B + 1$ | 0.96 | 19,541/19,541<br>100% | 19,541/19,541<br>100% | 0/19,541<br>0% | 0/19,541<br>0% | 19,541/19,541<br>100% |
| $\hat{B} = B + 2$ | 0.96 | 19,541/19,541<br>100% | 19,541/19,541<br>100% | 0/19,541<br>0% | 0/19,541<br>0% | 0/19,541<br>0% |
| $\hat{B} = B + 3$ | 0.96 | 19,541/19,541<br>100% | 19,541/19,541<br>100% | 0/19,541<br>0% | 0/19,541<br>0% | 0/19,541<br>0% |

**Table 6:** Output structures returned by the software.

(a) Fields of the signal output structure `signals`.
| Field | Type | Description |
| --- | --- | --- |
| I, II, III, aVR, aVL, aVF, V1–V6 | double | Recovered waveforms for each of the twelve standard ECG leads (one lead per named field). |
| units | char | Amplitude units of the lead signals (‘mV’, or ‘ADC’). |
| totalClipping | double | Total number of clipped samples across all leads. |
| hz | double | Sampling rate in Hz. |
| certified | double | Flag for if passed (1) or failed (0) certification. |
| *Optional fields if a rhythm strip is present* |  |  |
| rhythm | structure | Recovered waveforms for any rhythm strips. Each rhythm strip is associated with a field of its name (e.g. `rhythm.II`). |
| totalClippingRhythm | double | Total number of clipped samples across all rhythm strips. |

(b) Fields of the information output structure `info`.
| Field | Type | Description |
| --- | --- | --- |
| hz | double | Sampling rate in Hz. |
| leadOrder | cell | Lead names in the order used throughout the output. |
| leadsMode | char | Mode for augmented leads (‘calcleads’ or ‘pdfleads’). |
| ecgFormat | char | Page layout of the source ECG (e.g. ‘12×1’). |
| ecgFormatStr | char | Additional description of the PDF layout. |
| numPolylines | double | Total number of vector polylines extracted. |
| numLeadsDetected | double | Number of ECG leads detected. |
| cmMatrix | double | Affine transform mapping source coordinates to centimeters. |
| certified | double | Flag for if passed (1) or failed (0) certification. |
| diagnostics | struct | Structure containing results of validation tests. |
| lowPassFreq | double | Low-pass filter cut-off frequency in Hz. |
| lowPassFreq | double | Low-pass filter cut-off frequency in Hz. |
| XMLmicrovoltsLSB | double | $\mu\text{V}$ per ADU (assumed from MUSE) |
| unitsPerMv | double | Stream units per millivolt. |
| mmPerMv | double | ECG amplitude gain (mm per mV). |
| mmPerSec | double | ECG sweep speed (mm per second). |
| alpha | double | Stream-to-ADU conversion, $\alpha$. |
| offsets | double | Per-lead baseline offset for each lead in stream units. |
| numPages | double | Number of pages in the source PDF. |
| numStreams | double | Number of data streams. |
| clipping | struct | Nested structure with per-lead clipping details. |
| maxStreamUnits | double | Per-lead maximum value in stream units. |
| minStreamUnits | double | Per-lead minimum value in stream units. |
| *Optional fields if a rhythm strip is present* |  |  |
| rhythmStripLeads | cell | Rhythm strip lead names |

### 3.7. Validation of Python Implementation

The Python implementation (tested on Python 3.14.3 with numpy 2.4.5) of pdf2ecg was validated on all 29,152 PDFs including all format/gain combinations in both “calcleads” and “pdfleads” modes. Reconstructed waveform ADU values agreed exactly with the original MATLAB implementation in 100% of cases for all leads and rhythm strips. There was likewise 100% agreement on all derived calibration parameters, reconstruction metadata, and per-lead certification diagnostics.

## 4. Discussion

We present open source software (available in MATLAB and Python) for bit-exact reconstruction of ECG waveforms from MUSE ECG PDFs, utilizing vector data contained within the PDF rather than scanning and digitization of a rasterized image. Using this method, which utilizes the double quantization of the originally acquired raw waveforms which are used to generate the PDF, reconstructed PDF ECG waveform data are identical to the raw data exported directly from the MUSE ECG database. The software identifies waveform clipping which can prevent bit-exact reconstruction. Additionally, as the PDF does not explicitly encode all of the parameters needed to convert stream units to bit-exact ADUs and physical units or each lead’s baseline offset, we developed certification tests to ensure that the parameters being used are correct and waveform reconstruction is truly bit-exact. The software successfully reconstructed over 376,000 ECG leads without any processing errors, and 100% of leads without clipping were reconstructed bit-exactly. Tests to certify reconstructed waveforms as bit-exact functioned appropriately, with 0% of non-bit-exact reconstructed waveforms passing certification in sensitivity analyses.

The process of reconstructing ECG waveforms from a PDF with vector graphics has been previously explored [9–12], although our method and software provide important improvements. Previous iterations of PDF waveform reconstruction reported very low MAE of ~ 0.26*μ*V [9], which is similar to our reported value of ~ 0.25*μ*V, and which suggests that the majority of the error in both studies was due to the expected rounding errors associated with direct stream-to-mV conversion. The slightly higher error observed by Shie et al. may also be due to clipping which was not quantified in their analysis. Our method, in contrast, reconstructs the original ADUs which were used to encode the PDF stream unit data, resulting in bit-exact reconstruction with no error.

A benefit of using bit-exact reconstruction is that although sub-*μ*V errors due to rounding may have minimal effect on many downstream uses of ECG waveform data, it is not possible to definitively know if these small errors have negligible effects on every possible downstream use. Directly comparing any number of downstream outputs when using direct stream-to-mV conversion (with its low level errors), or bit-exact ADU reconstruction would not resolve this question, because any findings would be specific to the models tested. For example, showing a specific artificial intelligence ECG or heuristic annotation model has very similar outputs with bit-exact compared to approximate direct stream-to-mV reconstruction would not mean that the small differences might not cross a boundary threshold in a different model and produce a substantially different output. It is simply not possible to test all possible models. Bit-exact reconstruction, however, guarantees that *every* downstream use of the reconstructed data will be exactly the same as if the original raw data was used. This has significant implications for use of automated ECG analyses and artificial intelligence models, where it is not always possible to know how small changes in ECG waveforms might change model outputs. Using direct stream-to-mV conversion, also introduces errors that scale with the gain of the PDF. Therefore, the same exact ECG data, displayed in the same format at 2 different gains, will have different errors, and may give different downstream results. Our method of reconstructing the original ADUs avoids this inconsistency.

A second novel and critical contribution of our software is, after excluding clipping, the ability to certify the signal reconstruction as bit-exact without needing to confirm this with a paired raw data file such as XML. Unlike other raw digital ECG formats, such as XML or DICOM, where all of the relevant baseline offsets and conversion factors are directly available in the data file, PDFs do not directly provide the same level of data checking and validation as they are designed for data display and not data analysis. Although our method for determining variables such as voltage gain, lead offset, and the value of the mV-to-ADU ratio are automatic and robust with multiple levels of error checking during data extraction, without the source ground truth data or the true values of these parameters, a subtle change in how MUSE encodes PDF data in different versions could silently result in complete loss of bit-exact reconstruction and large reconstruction errors. Our certification tests, however, protect against this possibility. If past or future MUSE PDFs were/are encoded with a different value of *α*, or if the baseline offsets no longer align with the calibration signal baselines as we assume (due to a different *B*), the certification tests are able to detect inconsistencies by analyzing features of the PDF stream unit quantization. For this reason, our method is stronger than simply providing bit-exact waveform reconstruction, because we can provide certification that bit-exact reconstruction was performed without needing any ground truth signals to verify that this is true. When analyzing PDF-only data without any raw data to validate correct reconstruction, being able to know that reconstruction from the PDF is accurate is critical.

In addition to providing bit-exact reconstruction, while other published PDF ECG reconstruction programs only were tested/validated on a single PDF layout and gain, our software has been validated on 5 different PDF layouts, each at 3 voltage gains, and identifies and processes rhythm strips without needing any user input.

We also have characterized the prevalence of waveform clipping, which has not been a topic of much interest in the ECG digitization literature. Our software identifies leads with clipping so they are not silently propagated into downstream analyses. Clipping is highly dependent on PDF layout and gain, and is a critical issue when the goal is to faithfully reconstruct waveform data from an ECG PDF, as clipped data is lost and can never be recovered. Although a large proportion of clipped leads were due to artifact rather than physiological data being clipped outside of the PDF rendering bounds, the errors associated with clipping can be large and are not predictable. In many cases, small regions of clipping may not be important for downstream use, but in other cases, such as R peak detection, loss of an R peak could result in significant differences when the PDF reconstructed data is compared to XML data. Clipping potentially affects any ECG digitization method, including digitization of rasterized images, although detecting small regions of clipping on a digitized image is extremely challenging.

There has been a great deal of interest in ECG image digitization in recent years, with multiple groups developing both open-source and proprietary methods for ECG image digitization. It should be acknowledged that our method of PDF waveform reconstruction is *not* a replacement for the ability to digitize ECG images in general. Our method has a significantly narrower use case, and currently only works when a MUSE generated PDF with vector graphics encoding the ECG waveforms is available as a digital file. Printed, scanned, or photographed PDF ECGs cannot be reconstructed by our method. However, our described method is complementary to existing ECG image digitizers. If a MUSE ECG PDF is available, converting it into a rasterized image prior digitization is sub-optimal; state of the art image digitizers have an ~ 20 − 100 *μ*V RMSE [1–8], which can be completely eliminated with our method. Additionally, our PDF to waveform reconstruction pipeline is completely deterministic and analytical; there is no need for image rasterization, segmentation, thresholding, detecting artifact, assessing image compression, dealing with overlapping signals, feature extraction, or the “black box” of deep learning models.

Limitations of our software include that the current iteration only works with GE MUSE PDFs that are encoded with vector graphics. We did not have a large sample of other manufacturers’ ECG PDFs to validate the method on PDF files created outside of GE MUSE, but limited preliminary analyses show that Philips and Mortara ECG PDFs can also have their waveforms extracted in a similar way; it is simply a matter of determining how the signals are stored in the data stream(s), determining the conversion between stream units and physical units, and obtaining the correct baseline offsets. The current MUSE-only program is a strong backbone for adapting the presented method/software to other ECG manufacturers. Given that the data structure of ECG PDFs from different manufacturers will vary, it will be important to validate bit-exact reconstruction of other manufacturer’s PDFs, and our certification tests will be useful as the presented method and software is extended to other ECG manufacturers. We have provided open-source software in MATLAB and Python (http://github.com/BIVectors/pdf2ECG) under the GNU General Public License version 3 so any researcher can use, modify, and improve our software.

A second limitation is that although 12 × 1 format PDFs contain 10 seconds of all 12 ECG leads and therefore are completely equivalent to use of XML for any downstream data processing, all other formats do not contain all 12 leads for 10 synchronous seconds. Therefore use of the PDF in formats other than 12×1 is not directly the same as having access to the raw data in XML format. However, this issue is present for any image digitization program; it is not possible to digitize what is not on the page. Our method therefore does not get around the limitation that a PDF may not contain all of the data that is needed for a model that accepts XML-derived data of 12 10-second waveforms. Some digitization programs include methods of approximating the missing data [7], but how such generative methods perform on data reconstructed with our software is unclear and beyond the scope of this analysis.

Finally, in rare situations a PDF may have different filtering (such as a different low-pass filtering cutoff) compared to the raw acquired ECG data. In these cases the PDF’s filtering is applied to the reconstructed data, and there could be differences between the PDF reconstructed waveforms and use of raw ECG data or a separate PDF with different filtering applied. However, the reconstructed waveforms will be exact based on the filtering applied at the time of PDF encoding.

## 5. Conclusion

We present methods and open-source software for bit-exact ECG waveform reconstruction from 12-lead ECG PDFs generated by the GE MUSE system. Additionally, the method/software analyzes the encoded and quantized PDF data to provide certification that the reconstructed waveforms are bit-exact without need for a separate ground-truth data source. Although this method does not replace the need for general ECG image digitization, if MUSE generated PDFs are available, the presented method/software offers the ability to perfectly reconstruct the original ECG waveforms with no error, to certify that bit-exact reconstruction was achieved, and is therefore preferable to first rasterizing the PDF image and then using an image-based ECG digitizer. Extension to other manufacterers’ ECG PDFs is planned. Software source code is available in MATLAB and Python at http://github.com/BIVectors/pdf2ECG under the GNU General Public License version 3.

## Data Availability

The software described in this manuscript is available at http://github.com/BIVectors/pdf2ecg. Due to ethical and legal restrictions, the remaining datasets used in this study are not publicly available. The results underlying the figures and tables in this manuscript will be made available by the authors upon request, where possible

## 6. Author Roles

CRediT authorship contribution statement

Jonathan W. Waks: Conceptualization, Data curation, Formal analysis, Methodology, Resources, Software, Validation, Visualization, Writing – original draft, Writing – review and editing. Hans F. Stabenau: Writing – review and editing. Khaled Abdelrahman: Writing – review and editing. Daniel B. Kramer: Writing – review and editing. Arunashis Sau: Writing – review and editing. Nicholas S. Peters: Writing – review and editing. Fu Siong Ng: Writing – review and editing. Boroumand Zeidaabadi: Writing – review and editing. Konstantinos Patlatzoglou: Writing – review and editing. Libor Pastika: Writing – review and editing.

## Supplemental Methods and Results

### S.1. Example of Parsing Stream Data into Polylines

As an example, consider the following hypothetical data in a PDF data stream:

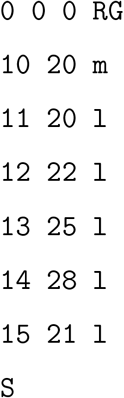

The first three `0` numeric tokens/operands are pushed into the operand stack. The parser then sees the alphabetic token `RG` which consumes the prior three operands (`0 0 0`) and sets the stroke color to black based on the RGB triplet `(0,0,0)`. Next the numeric tokens `10` and `20` are pushed into the operand stack, followed by the alphabetic operator `m` which starts a new polyline at point (`10,20`). Next operands `11` and `20` are pushed into the operand stack, followed by the operator `l` which adds the point (`11,20`) to the polyline. This continues for the additional points marked by operator `l` so that the polyline has 6 points at the time the operator `S` finalizes the polyline with color black. Each ECG lead waveform on the PDF has its own polyline created in this way.

### S.2. ADU to Stream Unit Relationship

The relationship between stream units and mV is best obtained from the minor grid lines as noted in Section 2.6. In theory it can also be obtained from information explicitly contained within the PDF as outlined below. However, we **do not** recommend use of this method as it has inherent inaccuracies built into it. We are presenting this alternative method here to illustrate why it is not ideal, as we initially thought this method was superior to using PDF lines to determine the relationship between stream units and mV. In reality, the error in 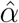 generated by this method will fail certification, although it is not large enough to prevent bit-exact reconstruction for most ECGs (see Supplement Section S.6)

The conversion between stream units and ADUs (*α*) is supplied by 4 conversions within the PDF file: the content stream’s current transformation matrix (cm) which is encoded as `a b c d e f cm` in the decompressed content stream [15] (see Section 2.4), which converts PDF stream units to PDF points, the PDF specification’s fixed point-to-mm constant (1 point = 1/72 inch = (25.4/72) mm), the ECG calibration printed on the document (e.g. 10 mm/mV), and the known conversion factor from mV to ADUs (4.88 *μ*V per ADU).

In parsed stream coordinates, the first coordinate (*x*) corresponds to time (*t*) and the second coordinate (*y*) corresponds to voltage (*v*). The cm matrix is a 6-number affine matrix defined as [15]:

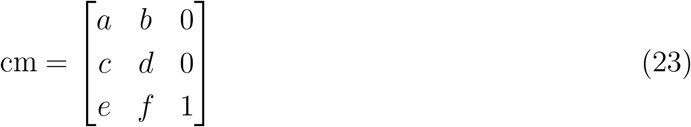

which transforms (*x*_stream_, *y*_stream_) in stream coordinates to points in PDF space (*x*_pt_, *y*_pt_) via the following equations [15]:

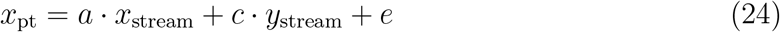

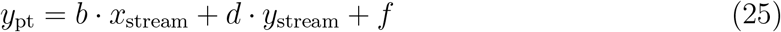

MUSE-generated PDFs use a transformation of the form [*a b c d e f*] = [0 *s* −*s* 0 *e* 0] with *s* = 0.02835 points/stream unit, *e* = 612 points (the 8.5-inch portrait page width), and *a* = *d* = *f* = 0. The opposite signs of *b* and *c* encode a 90 deg rotation (since |*b*| = |*c*|, a single scalar, *s*, is used), and the *e* = 612 translation places the rotated content within the page bounds; together they orient the ECG correctly on the portrait page, which the viewer then rotates 90 deg to landscape for viewing/display. Because the rotation and translation affect only where the rendered marks appear on the page and not the encoded signal values, only the magnitude *s* is needed for signal recovery. The scalar *s* converts stream units to PDF points, and the PDF specification’s point-to-mm constant ((25.4/72) mm per point) then converts points to mm.

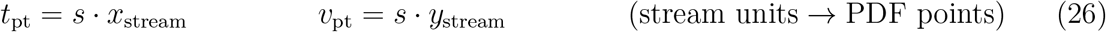

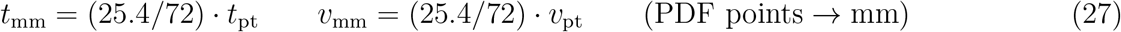

The decompressed stream is then searched for information on the time and voltage calibration by looking for the characters “25 mm/s” and *G* “mm/mV” where *G* is an integer extracted by regular expressions (see Methods Section 2.5). Combining the stream-to-point, point-to-mm conversion, and extracted ECG calibration, gives the per-axis conversion factors assuming a sweep speed of *T* mm/s and a voltage gain of *G* mm/mV:

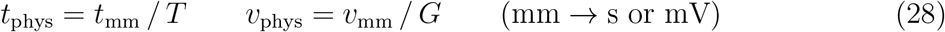

Using the nominal values of *T* = 25 mm/s and combining Eqs. (26–28):

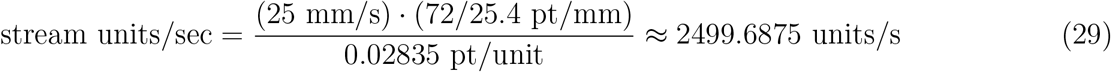

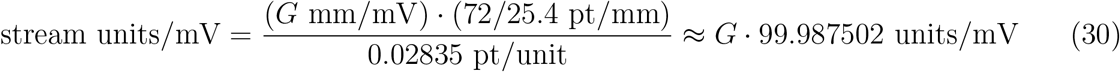

and therefore:

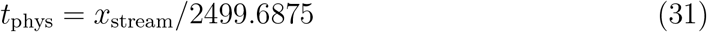

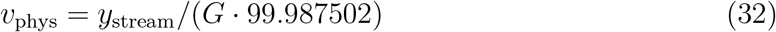

Written differently, we see that at a gain of 10 mm/mV, because 1 mV = 999.87502 stream units, it follows that:

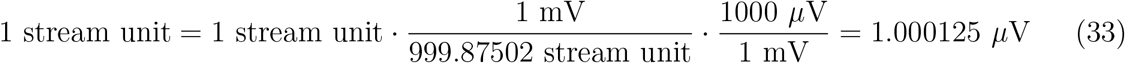

and since 1 ADC unit = 4.88 *μ*V:

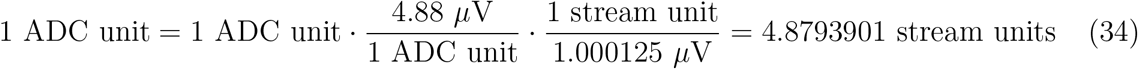

For any *G* units/mV, and taking *s* = 0.02835 pt/unit from the cm matrix, we define the ratio of stream to ADC units, *α*, as:

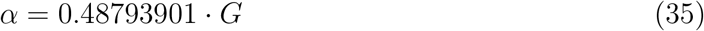

As noted in the main Methods Section 2.6, these relationships are slightly incorrect. The true relationship between stream units and physical units is 1 mV = 1000 stream units and *α* = 0.488 · *G*. Tracing the internal conversion pathway results in 1 mV = 999.87502 stream units because the value of *s* in the cm matrix is truncated to 5 decimal places: *s* = 0.02835. The value of *s* which would be required to obtain a conversion of 1 mV = 1000 stream units is 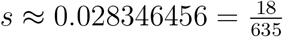. Although this difference has no perceptible effect on how the PDF is rendered, and would have a trivial effect on most direct stream units to mV conversions, it does result in a slightly incorrect value of the stream-to-ADU conversion, *α*, at larger ADU values the small error compounds, and it can have some downstream effects during conversion back to bit-exact ADUs. See Supplement Sections S.5 and S.6 for more details.

### S.3. Derivation of Allowed Values of Δ*S*

Consider *Y* as the un-rounded value of ADU values (*A*) multiplied by *α* with offset *B*. When *Y* is rounded, we get stream units, *S*, which are plotted in the PDF. We will look at Δ*S*_*k*_ which is the change in stream units for a *k*–unit change in integer ADU values between subsequent ADU samples, so *A*_*n*+1_ − *A*_*n*_ = *k*. We have:

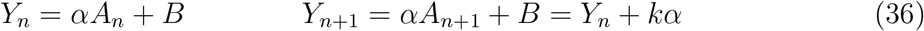

Stream units are the rounded values of *Y*, so

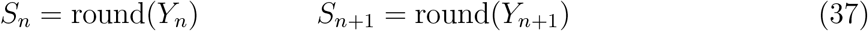

and their difference, Δ*S*_*k*_ is:

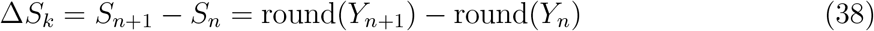

Because *Y*_*n*+1_ = *Y*_*n*_ + *kα*:

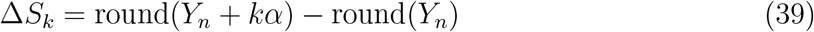

We then define the rounding error of sample *n* as:

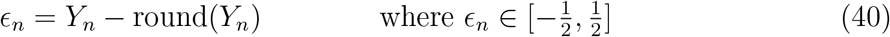

and after rearranging so that *Y*_*n*_ = *ϵ*_*n*_ + round(*Y*_*n*_), and substituting back into Eq 39:

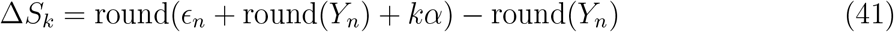

round(*Y*_*n*_) is just *S*_*n*_ by definition, so:

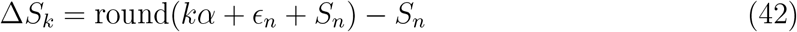

*S*_*n*_ is an integer, and it can therefore be pulled outside of the rounding operation:

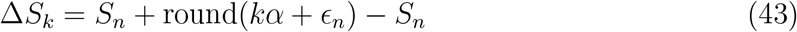

The *S*_*n*_ cancel, and therefore we finally have:

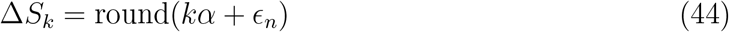

Since 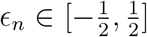, 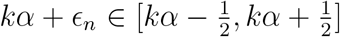, and therefore, after rounding, Δ*S*_*k*_ can only be the two integer values bracketing *kα*, namely ⌊*kα*⌋ and ⌈*kα*⌉. For a unit step (*k* = 1) and *α* = 4.88, this gives Δ*S*_1_ = 4 or 5, with the relative frequency of the two values determined by the fractional part of *kα*. See Figure 4 for further details. Note that although Δ*S*_*k*_ involves the rounding of two samples, it depends on only a single rounding error *ϵ*_*n*_; because consecutive positions differ by the exact quantity *kα*, the second sample’s rounding is fully determined once *ϵ*_*n*_ is known/fixed.

### S.4. MUSE Rounding of the Augmented Leads

During validation of the augmented limb leads (aVR, aVL, aVF), we observed that recovered values differed from the reference values by up to one-half ADU. Analysis of the encoded stream’s first-difference spectrum (Figure 2) showed steps at integer multiples of *α* with no half-*α* steps, indicating that MUSE rounds the augmented leads to integer ADU values prior to stream encoding, rather than preserving the native half-ADU value precision. To identify the rounding convention, we compared the recovered integers in leads aVR, aVL, aVF against their ground truth values (Eq. 15) under different rounding conventions, and round-half-toward-zero reproduced the recovered stream exactly in all leads, over thousands of tested leads.

In layouts where leads I and II are displayed simultaneously with the augmented leads, direct reconstruction of the augmented leads in native half-ADU values is possible. In 3×4 layouts, however, the augmented leads and leads I and II are not simultaneous, precluding this reconstruction. Reconstruction of the augmented leads in 3 × 4 layouts is therefore limited to MUSE’s pre-encoding rounded integers, which we recover bit-exactly. The residual discrepancy of the rounded values relative to the native half-ADU values is therefore a property of how MUSE internally calculates the augmented leads and is not indicative of failure of the reconstruction method. Given that the raw data in XML format does not include the augmented leads, there is no “correct” ground truth for the augmented leads, and the proposed software allows bit-exact recovery of either definition for all formats except 3 × 4 where bit-exact recovery is only available for the rounded values which are used for PDF generation.

### S.5. Bit-exact Certification Test Derivations

For the following derivations we will use the following notation: *α* and *B* are the values used by MUSE at the time of encoding from raw ADUs to PDF stream units, and 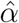 and 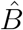 are the values that are used by the software for waveform reconstruction. Because the PDF does not explicitly encode the values of *α* and *B*, the certifications tests are designed to determine if 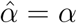 and/or 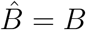, and if this is true we are guaranteed to have bit-exact reconstruction. If 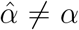 and/or 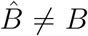, then bit-exact reconstruction may still occur, but we cannot guarantee that this will be true.

#### S.5.1. Residue Width Test, (*W*)

##### S.5.1.1. Residue Width Geometry

Under the encoding model, we assume 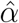, which is the value of the stream-to-ADU conversion ratio which we use, is the same value as was used to encode the PDF stream values which is *α*. The true encoding scale, *α*, is not directly observed. We therefore need a test to certify that 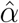 is consistent with *α*.

The encoding relationship which generates stream units from ADUs, as previously defined in Eq. 10, uses *α*, and is:

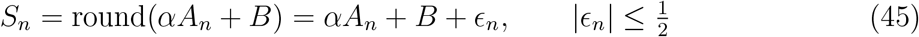

We define the error between 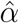 and *α* as *δ*_*α*_:

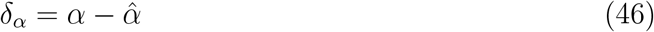

substitute 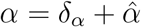 into Eq. 45:

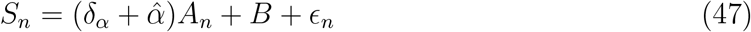

and then take modulo 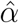 of both sides of the equation:

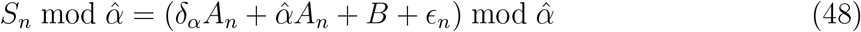

This has the effect of wrapping the stream units around a circle of circumference 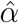 and is illustrated in Supplemental Figure 2. Taking mod 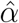 of both sides of Eq. 48 makes the 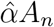 term vanish, because *A*_*n*_ is always an integer, and 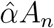 is therefore an integer multiple of 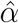 which reduces to 0 mod 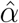. As a result:

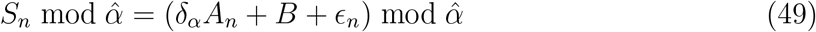

We first examine what happens when 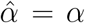 and *δ*_*α*_ = 0. In this case, the remaining *δ*_*α*_*A*_*n*_ term in Eq. 49 also vanishes, leaving:

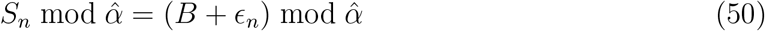

*B* is a constant, and 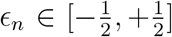, so the residues collapse into a circular arc centered on *B* mod 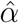, extending 0.5 stream unit in either direction (the modulo operator handles any wrapping around the circle, so this is not the same as a linear interval). As a result, if *δ*_*α*_ = 0, the length of the residue arc is determined entirely by the range of *ϵ*_*n*_ and is therefore always ≤ 1 stream unit.

##### S.5.1.2. Definition of W

We do not know the values of *ϵ*_*n*_, but we do know the values of *S*_*n*_ and 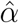, and we therefore work with the left side of Eq. 50. The residues (*r*_*n*_) are obtained directly from the stream units and 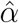 as:

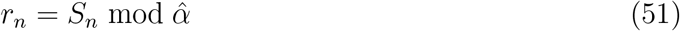

We define *W* as the width of the shortest circular arc containing all of the observed residues, *r*_*n*_. To find the value of *W* we sort the *N* unique values of *r*_*n*_:

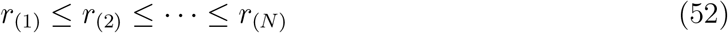

and find the gaps between sorted residues, 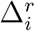:

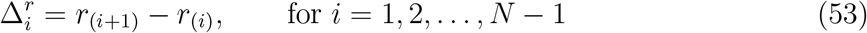

also accounting for the wraparound gap (with circle of circumference 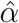):

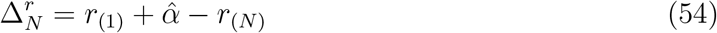

We then find the largest gap, max 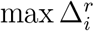. As the circle has circumference 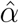, 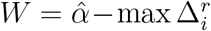 (see Supplemental Figure 3). If *δ*_*α*_ = 0, because the residues therefore span at most 1 stream unit of circular arc, it follows that *W* ≤ 1.

##### S.5.1.3. Quantization of W

*W* can be *<* 1 despite 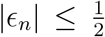 due to quantization; just as the values of *S*_*n*_ are not continuous, the residues of *S*_*n*_ mod 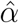 are also not continuous, and therefore they may not occupy every value on the 1 stream unit residue arc.

Because 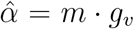 · *G* (Eq. 4) is a product of the assumed stream-to-ADU conversion, *m*, with two integers read from the PDF (*g*_*v*_ and *G*), it is a rational number which can be written in lowest terms as:

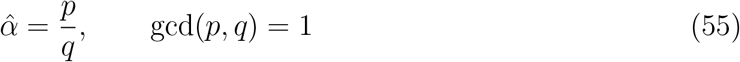

Both *p* and *q* are determined by the value of 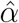 and are fixed before any testing is performed. For all MUSE PDFs evaluated in this study, 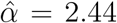, 4.88, and 9.76 stream units/ADU at gains of 5, 10, and 20 mm/mV, corresponding to *q* = 25 with *p* = 61, 122, and 244, respectively. We can therefore re-write the encoding equation (Eq. 45) using 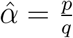 and then multiply each side by *q*:

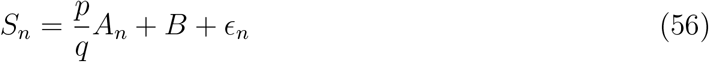

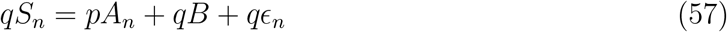

We then can rearrange Eq. 57 in terms of *qϵ*_*n*_:

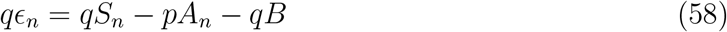

Because *S*_*n*_, *A*_*n*_, *B, p*, and *q* are all integers, every term in Eq. 58, including *qϵ*_*n*_, must also be an integer.

We define integer *j*_*n*_ = *qϵ*_*n*_. As a result:

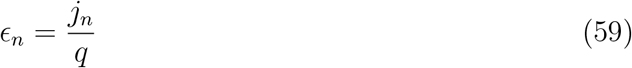

Because 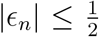 and *q* is an integer, *j*_*n*_ must be a sequence of integers that span between 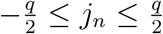. For odd *q*, 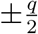 is never an integer, exact half-unit rounding ties never occur, and *j*_*n*_ can take exactly *q* values.

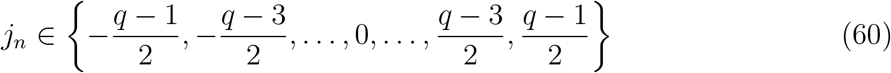

and from Eq. 59 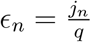, so:

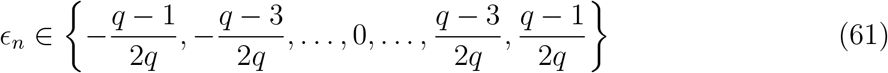

Because of this quantization, the bound for *ϵ*_*n*_ is actually smaller than 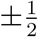 since the value of 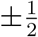 can never be reached. The maximum width of the *quanitized* residues in the case where *δ*_*α*_ = 0 is therefore:

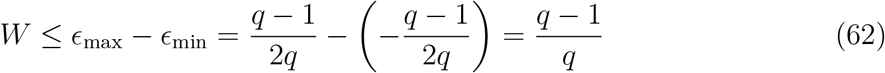

and therefore when *δ*_*α*_ = 0:

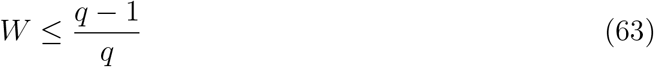

For even *q*, both endpoints of 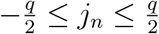 are integers and whether both are included in the interval depends on MUSE’s tie-breaking convention, which is not observable from the PDF; for this reason, if *q* is even, the software then applies the conservative bound *W* ≤ 1. This case does not arise for any 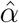 encountered with MUSE generated PDFs, but could appear in the future if the value of *α* were to change. As *q* gets larger (as 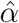 contains more decimal points) (*q* − 1)*/q* →1. In that case the quantized bound of Eq. 63 approaches the generic bound *W* ≤ 1 and provides little additional discrimination. The software reports *q* so that this loss of discrimination in the value of *W* can be quantified.

##### S.5.1.4. Quantization of Residues in MUSE ECGs

For *q* = 25, which is present for all MUSE ECGs regardless of gain, based on Eqs. 60 and 61 we obtain:

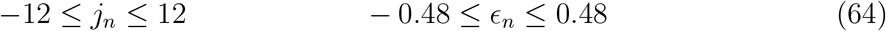

As a result, instead of using the limit of *W* ≤ 1, in all MUSE ECG PDFs, due to quantization of the residues with *q* = 25, (*q* − 1)*/q* = 0.96, and, when *δ*_*α*_ = 0:

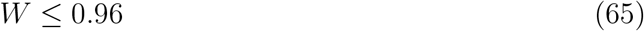

This is confirmed in the processed PDF data as shown in Table 5.

##### S.5.1.5. Failure of the Residue Width Test

We now return to Eq. 49 when *δ*_*α*_ ≠ 0, and therefore 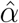 is *not* consistent with *α*. In this case, the *δ*_*α*_*A*_*n*_ term does *not* vanish mod 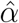, and the residues are therefore no longer constrained by the rounding error *ϵ*_*n*_ alone; the residues now also vary with *A*_*n*_ through the *δ*_*α*_*A*_*n*_ term. As *A*_*n*_ varies across the range of values contained within an ECG lead, the *δ*_*α*_*A*_*n*_ term can spread the residues outside of the bound established when *δ*_*α*_ = 0, and therefore *W >* (*q* − 1)*/q* (see main Methods Section 2.12, Figure 6, and Supplemental Figure 2). The residue width test can also fail due to clipping even if 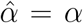, although clipping would be identified separately

##### S.5.1.6. Avoiding Floating Point Errors

To evaluate if *W* ≤ (*q* − 1)*/q*, we have to find the locations of the residues by evaluating *r*_*n*_ = *S*_*n*_ mod 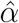. Taking an integer mod 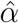, can introduce floating-point errors that can exceed the margin of Eq. 63 (e.g. a value of 0.96000000001 will fail the test). To avoid these floating-point errors, we return to Eq. 57, and note that the *A*_*n*_ term is now multiplied by integer *p* instead of non-integer 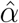. We therefore repeat the prior procedure: to make the *A*_*n*_ term vanish we take mod *p* of both sides of Eq. 57 and define the value of the resulting residues as *R*_*n*_:

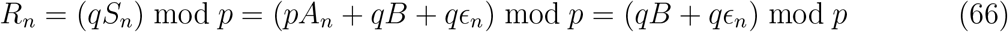

Note how Eq. 66 is just Eq. 50 with each side of the equation multipled by *q*. As *S*_*n*_, *q*, and *p* are integers, *R*_*n*_ = *qS*_*n*_ mod *p* must also be an integer with range 0 ≤ *R*_*n*_ ≤ *p* − 1. The residues *R*_*n*_, which are now obtained by taking (*qS*_*n*_) mod *p* instead of *S*_*n*_ mod 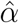, therefore occupy *p* discrete positions on a circle of circumference *p*, rather than values on a circle of circumference 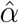.

Because 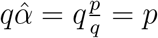, multiplying by *q* effectively maps the residue circle of circumference 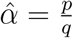 onto a circle of integer circumference *p*. From Eqs. 51 and 66, the two sets of residues are related by *R*_*n*_ = *q r*_*n*_, and therefore:

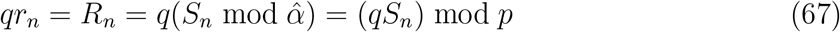

Therefore, every residue position *R*_*n*_, and therefore every distance between the *R*_*n*_ residues 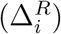, is multiplied by exactly *q* when compared to *r*_*n*_. As a result, if 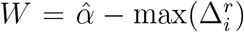, since 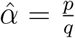, then 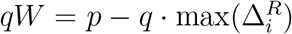. The shortest arc containing all *R*_*n*_ therefore has width

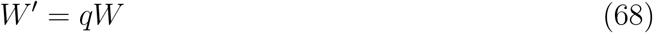

*W*^*′*^ is not measured in stream units, and is instead measured in quanta of 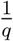 stream units (which we will call “residue units”). Therefore multiplying Eq. 63 (*W* ≤ (*q* − 1)*/q*) by *q* preserves the direction of the inequality, and the certification equation therefore becomes:

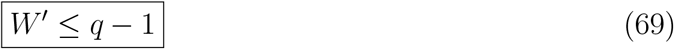

*W*^*′*^ is obtained exactly as *W* was: after ordering the unique values of *R*_*n*_ and calculating the gaps between successive values of *R*_*n*_ including the wraparound gap, the sum of all gaps, adds to *p*, which is the circle’s circumference. Therefore, to obtain the shortest arc containing all of the residues *R*_*n*_:

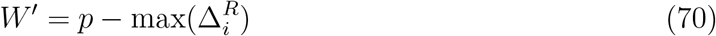

Because every quantity in Eqs. 66–70 is an integer, and their products remain well within the exactly representable range of double-precision arithmetic, the comparison is exact and no numerical tolerance is required. For this reason, *W* = (*q* −1)*/q* is never calculated; instead certification is evaluated exactly as *W*^*′*^ = *q* − 1, and *W* = *W*^*′*^*/q* is additionally reported for interpretability.

##### S.5.1.7. Applying The Residue Width Test

We therefore apply the residue width test by converting 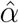 into its rational/fractional form 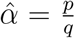, calculating *R*_*n*_ = (*qS*_*n*_) mod *p*, ordering *R*_*n*_, finding the maximum distance between all combinations of 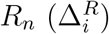, and then calculating 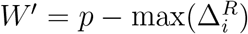 (Eq. 70). All values are in integers, so no floating-point issues can occur. The test is passed if *W*^*′*^ ≤ *q* − 1, and the test otherwise is failed. For all MUSE PDFs, *q* = 25 and the test is passed if *W*^*′*^ ≤ 24.

Passing *W*^*′*^ ≤ *q* − 1 indicates that the observed stream encoding (*S*_*n*_) is consistent with the proposed value of 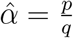 which was used to generate the stream unit values from ADUs; it does not, by itself, establish that 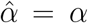 exactly. Conversely, failure of this test does not necessarily imply that bit-exact reconstruction has failed, because the residue width test is far more sensitive to errors in 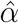 than bit-exactness requires, and a sufficiently small value of *δ*_*α*_ may not move any sample across an ADU rounding boundary. Failure, with *W*^*′*^ *> q* − 1 indicates that bit-exactness can no longer be certified from the residue structure, and that the user should verify the value of the encoding parameters (*m, g*_*v*_, *G*) from which 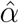 is derived. Further discussion about how much 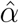 must deviate from *α* is available in Supplement Section S.6.

The residue width test result also depends on how much of the residue grid the waveform occupies. Leads that are extremely flat or disconnected have limited unique values of *A*_*n*_, and therefore the observed residues may not occupy the full range of possible residues as would be seen with a wider sampling of *A*_*n*_ values. As a result, a flat/disconnected lead may pass the residue width test even if *δ*_*α*_ ≠ 0 because a limited variety of *A*_*n*_ values results in a narrow distribution of *R*_*n*_. The software therefore reports the number of occupied residue classes alongside *W*, and if a single class is occupied, the test is indeterminate. Examples of the residue width test are shown in Figure 6A-D and Table 5.

#### S.5.2. Greatest Common Divisor (GCD) Test

The residue width test *W* (Eq. 69) has one important blind-spot. If 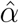 is an exact integer submultiple of *α*, so that 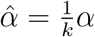 for integer *k >* 1, then 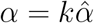 and

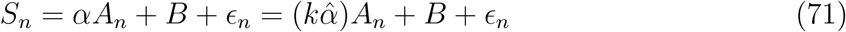

The 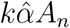 term remains an exact multiple of 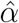 and vanishes under the mod 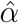 operation:

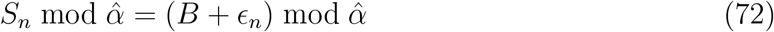

which is exactly the same as Eq. 50. *W*^*′*^ can therefore satisfy Eq. 69 even though 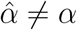 and the reconstructed values are scaled by an integer factor *k* (see Eq. 12):

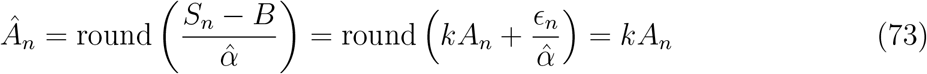

because 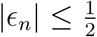 and 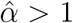, 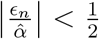 and is removed by the rounding operation. Note that this issue does *not* also hold for 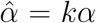 because 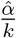 does not vanish mod 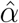 for *k >* 1. The requirement that *α* and 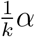 must be *>* 1, needed for distinct ADU values to map to distinct stream values, also bounds which submultiples are reachable. Since 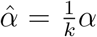, only *k < α* is possible. For *α* = 4.88, *k* is therefore restricted to values of 2, 3 and 4.

The submultiple ambiguity of the residue width test is resolved by using the greatest common divisor (GCD) of all non-zero sequential sample differences in the reconstructed signal, Δ*Â*_*n*_ = *Â*_*n*+1_ − *Â*_*n*_ (see Supplemental Figure 4 and Figure 7).

**Figure 7:**
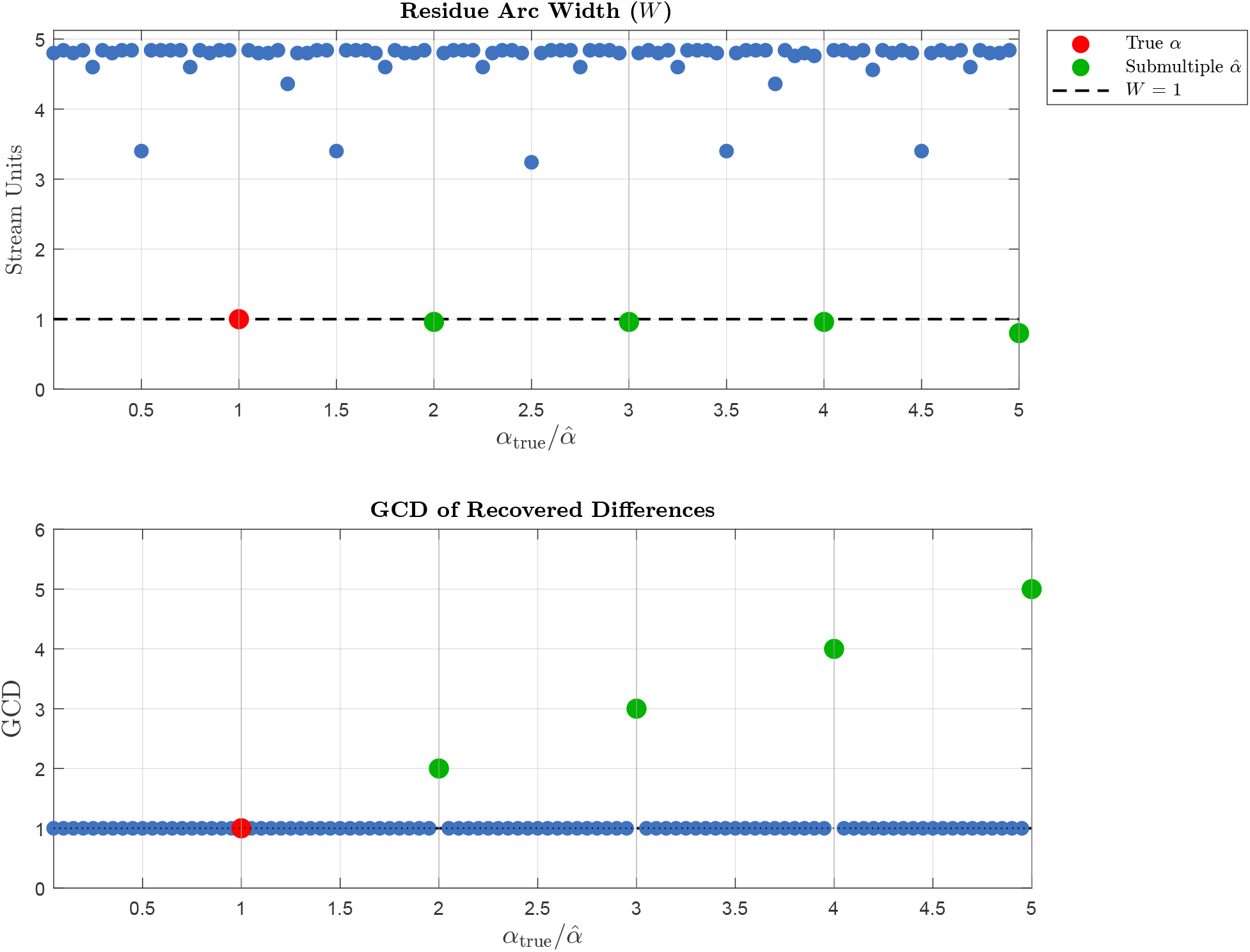
Summary of using residue width (*W*) and GCD of the recovered differences to determine certification of ECG waveforms where 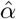 is an multiple or submultiple of *α*_true_. The top panel shows the value of *W* as a function of the ratio of 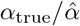. Higher number indicate 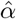 is getting smaller relative to *α*_true_. When 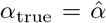 the ratio = 1, and the red dot shows that *W* ≤ 1 and GCD = 1. *W* ≤ 1 only when 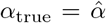 or when 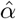 is an integer submultiple of *α*_true_ 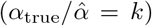, for integer *k*, as shown for the green points {2, 3, 4, 5}. Although an integer submultiple (green dots) passes the residue width test, as can be seen in the bottom panel, the GCD is *>* 1 and these values of 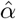 would therefore appropriately fail certification at the GCD test. For all integer multiples of *α*_true_, which occurs for 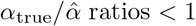, the residue width test (*W*) always fails and the GCD test is not required.

If no non-zero differences exist (such as a flat or disconnected lead), the GCD is undefined, the test is not assessable, and the software reports this. For 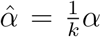, *Â*_*n*_ = *kA*_*n*_, Δ*Â*_*n*_ = *k*Δ*A*_*n*_, and therefore all non-zero reconstructed differences are divisible by integer *k*. A GCD of 1 implies that *k* = 1 and therefore excludes 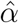 being an exact integer submultiple value of *α* used for encoding. If the GCD is *>* 1 the reconstruction fails certification. A GCD *>* 1 does not by itself prove that 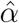 is a submultiple of *α*, since a signal whose true ADU values happen to change only in multiples of *k* would also produce the same result. This, however, would be vanishingly unlikely for a lead with many transitions, but it cannot be excluded when only a few non-zero differences are available. The software therefore reports the number of non-zero differences alongside the GCD. Across the 29,152 PDFs and *>* 357, 000 leads evaluated in this study, a GCD *>* 1 was never observed under the correct value of 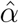. Figure 6D and Figure 7 illustrates how *W* and GCD together can certify bit-exact recovery for different combinations of 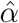 and *α*.

#### S.5.3. Baseline Residue Test

The baseline residue certification test is performed only after the proposed value of 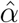 passes both the residue width and GCD tests, because the predicted residue window, as discussed below, is undefined if the value of 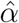 is incorrect. Because *B* is an integer PDF stream coordinate, the proposed baseline value 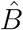 predicts an exact discrete set of allowable residues centered on a specific location.

From Eq. 66, assuming that 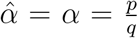, and using our obtained value for the offset 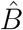, the observed residues must satisfy

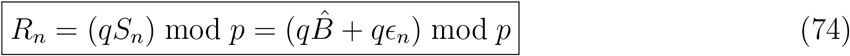

We therefore have 2 ways of calculating *R*_*n*_: we can calculate (*qS*_*n*_) mod *p* or 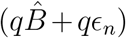 mod *p*. We don’t know the values of *ϵ*_*n*_, but we can bound the possible values of *R*_*n*_ using 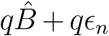 mod *p*. Since we have already determined that 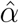 and therefore *p* and *q* are correct, and the stream units *S*_*n*_ are extracted directly from the PDF datastream, if the *R*_*n*_ calcualted from (*qS*_*n*_) mod *p* do not fall within the window/range defined by 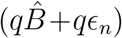 mod *p*, the value of 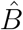 must be inconsistent with the true offset value *B*.

The baseline residue test therefore compares two independently derived values/ranges of *R*_*n*_ as noted in Eq. 74; 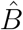 predicts a specific location and arc width of allowable residues centered on 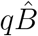 mod *p*, with width and allowed quantized residues determined by *qϵ*_*n*_, and without reference to any value of *S*_*n*_. This range must match the observed residue locations independently calculated from (*qS*_*n*_) mod *p* without any reference to *B*. If every *R*_*n*_ calculated from (*qS*_*n*_) mod *p* falls within the window calculated from 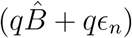 mod *p*, the assumed baseline 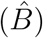 is consistent with the offset used for encoding (*B*). If *any R*_*n*_ falls outside of the window predicted by 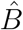, then 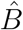 is not consistent with *B* under the assumed encoding model, and the waveform reconstruction fails certification.

An error of *δ*_*B*_ stream units in 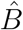 displaces the predicted window by *qδ* mod *p* residue units (where 1 residue unit is 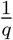 stream units). A one-stream-unit error therefore shifts the window by exactly *q* residue units, every observed residue then falls outside of the window predicted by 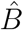, and the test fails. Larger baseline errors wrap around the circle and may partially overlap the observed residues. The test is therefore sensitive to small integer baseline errors that arise in practice from incorrect extraction of lead baselines. Because the window depends on 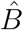only through 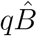 mod *p*, baselines differing by an exact multiple of *p* stream units are indistinguishable. Such an error would offset every reconstructed sample by the same integer multiple of *q* ADU while leaving all sample-to-sample differences unchanged. Such exact shifts in every lead are highly unlikely. Errors of any other magnitude displace the window and are detected. Examples of the baseline residue test are shown in Figure 6E-F.

#### S.5.4. Certification result

The three certification tests are applied hierarchically. The baseline-residue test is assessed only when the residue-width and GCD tests both pass. If an upstream test does not pass, the baseline is reported as not assessed rather than failed. A reconstruction is certified when: (1) the observed residue width is consistent with the used value of 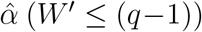, (2) the GCD of the nonzero reconstructed differences is 1, excluding exact integer-submultiple 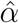, and (3) every observed integer residue lies within the discrete residue window predicted by the candidate baseline 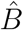. Together, these tests establish that the parameters required for bit-exact recovery are satisfied without reference to the source XML data.

### S.6. Tolerance of 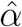 Error

As noted in Supplement Section S.5, *W*^*′*^ ≤ (*q* − 1) implies that 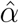 is consistent with the value of *α* used for PDF encoding. However, as noted, *W*^*′*^ *>* (*q* − 1) implies that bit-exact reconstruction can no longer be guaranteed, not that bit-exact reconstruction will necessarily fail. We will show how large an error in 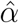 must be to cross a rounding boundary and lose bit-exact reconstruction. For this section we will assume that the value of *B* is correct (see Supplemental Section S.7 for information on how errors in *B* affect bit-exact reconstruction).

We start with the reconstruction equation (Eq. 10) and assume that our value of 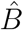 is correct 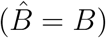:

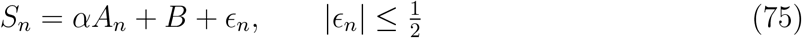

We define 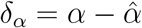 and substitute this into Eq. 75:

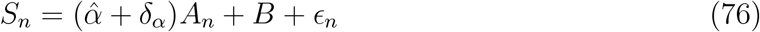

We then subtract *B*, divide both sides by 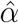, and round to obtain *Â*_*n*_:

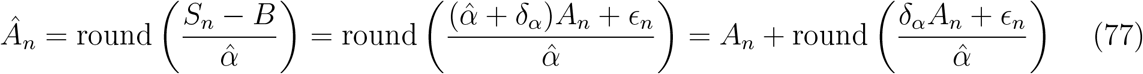

Therefore, we end up with *Â*_*n*_ being equal to the true value of *A*_*n*_ plus an error term 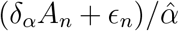. The full error term (which reduces to the familiar 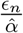 if *δ*_*α*_ = 0) must be *<* 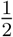 to be eliminated by the rounding operation:

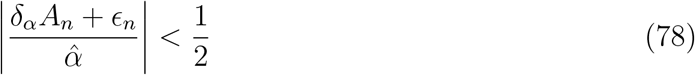

This therefore requires that

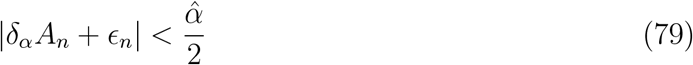

Because *ϵ*_*n*_ spans an interval of width 1, the quantity |*δ*_*α*_*A*_*n*_ + *ϵ*_*n*_| represents an interval 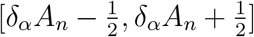. The upper edge of the interval 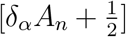 represents the values of |*δ*_*α*_| and |*A*_*n*_| which first cross the critical value of 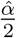, at which point only samples with *ϵ*_*n*_ near 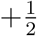 fail. Other samples are “rescued” by a negative *ϵ*_*n*_ which decreases the total error below 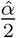 and allows bit-exact reconstruction to occur despite error in 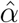. Therefore, these values represent the transition where bit-exact reconstruction for that combination of |*δ*_*α*_| and |*A*_*n*_| is no longer guaranteed, although it still may be possible.

To determine the maximum error |*δ*_*α*_| which will still allow bit-exact reconstruction for an entire ECG lead, the bound must hold true for all *A*_*n*_, and we therefore use the largest magnitude *A*_*n*_ which equals |*A*_max_|. Rearranging Eq. 79 in terms of |*δ*_*α*_| and using the worse case scenario where 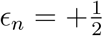:

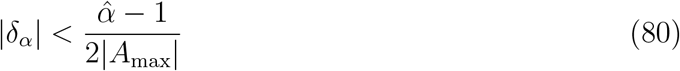

The maximum |*δ*_*α*_| which allows bit-exact reconstruction for the entire ECG lead therefore depends on the value of 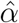 itself, and the actual ADU values which encode the waveform. Larger 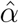, as occurs with larger gains, provides an additional buffer to guarantee bit-exact reconstruction, and signals with very large amplitude increase the risk of a fixed error in 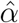 causing loss of bit-exact reconstruction (see Supplemental Figure 5).

The lower edge of the interval 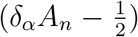 represents the values of |*δ*_*α*_| and |*A*_*n*_| where even the most favorable value of 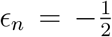 cannot reduce the total error below 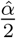. These values, therefore, represent the transition where bit-exact reconstruction is guaranteed to fail. Therefore, the minimum |*δ*_*α*_| which will result in failure of bit-exact reconstruction in at least 1 sample of a lead is:

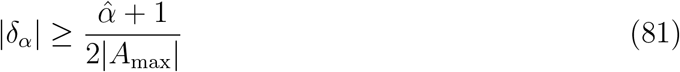

These 2 transitions are illustrated visually in Supplemental Figure 6. Similar to the upper bound, larger 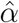 and smaller *A*_*n*_ increase the tolerance for error.

The two transition locations for |*δ*_*α*_| are set by 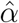 and depend on the product *δ*_*α*_*A*_*n*_. |*δ*_*α*_| and *A*_*n*_ trade off directly: the same failure occurs for a large error on a small ADU value as for a small error on a large ADU value. As a result, with a small error in 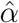as occurs when using the value of 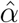 obtained by incorrectly using the internal PDF unit conversions as outlined in Supplement Section S.2, small number of extreme valued *A*_*n*_ could fail bit-exact reconstruction while the remainder of the waveform is accurately reconstructed (as shown in Supplemental Figure 5). The relationship between |*δ*_*α*_| and |*A*_*n*_| and the locations of bit-exact transitions is illustrated in Supplemental Figure 6.

### S.7. Tolerance of Offset (*B*) Error

The minimum error in 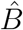 which is needed to cross a rounding boundary and break bit-exact reconstruction can also be calculated in a similar way to the calculation of errors in 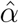 (see Supplemental Section S.6).

Similar to the analysis in *α* errors, we start with the reconstruction equation (Eq. 10) and assume that 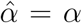 since the baseline certification test only applies if this is true and prior certification tests have passed.

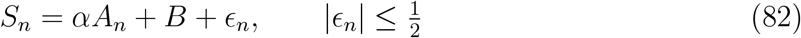

We then define 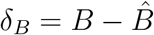 and substitute 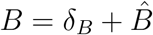 into Eq. 82:

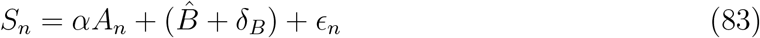

We subtract 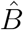, divide both sides by *α*, and round to obtain *Â*_*n*_:

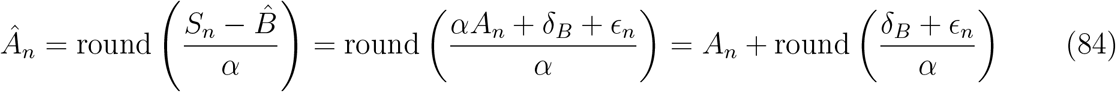

Therefore, we end up with *Â*_*n*_ being equal to the true value of *A*_*n*_ plus an error term (*δ*_*B*_ + *ϵ*_*n*_)*/α*. The full error term (which reduces to the familiar 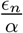 if *δ*_*B*_ = 0) must be 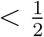 to be eliminated by the rounding operation:

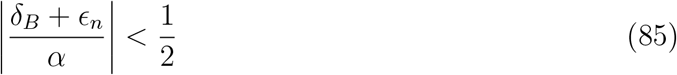

This therefore requires that

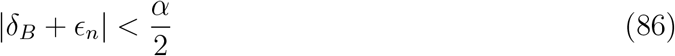

Because *ϵ*_*n*_ spans an interval of width 1, the quantity |*δ*_*B*_ + *ϵ*_*n*_| represents an interval 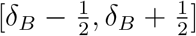. The upper edge of the interval 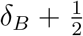 represents the value of |*δ*_*B*_| which first crosses the critical value of 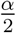, at which point only samples with *ϵ*_*n*_ near 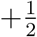 fail. Other samples are “rescued” by a negative *ϵ*_*n*_ which decreases the total error below 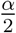 and allows bit-exact reconstruction. The error term for 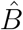 (Eq. 86) is constant (other than the rounding error), unlike the error term for *α* (Eq. 79) which depends on the value of *A*_*n*_ in addition to the value of the error in *α*.

To determine the maximum error in |*δ*_*B*_| which will still allow bit-exact reconstruction, we rearrange Eq. 86 in terms of |*δ*_*B*_| and use the worse case scenario where 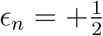:

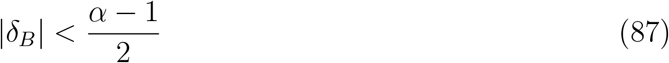

Values of *δ*_*B*_ below this threshold, which is determined only by *α*, are guaranteed to be bit exact.

We then consider the lower edge of the interval 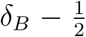, which represents the value of |*δ*_*B*_| where even the most favorable value of 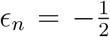 cannot reduce the total error below 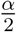. Therefore, once this threshold is crossed, every sample will fail bit-exact reconstruction and be inaccurate. Therefore the minimum value of |*δ*_*B*_| for which bit-exact extraction will always fail is given by:

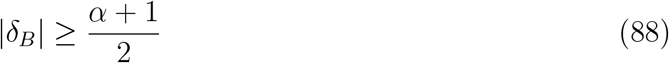

The effect of *δ*_*B*_ is illustrated in Supplemental Figures 7 and 8. Unlike the error in *α*, the bit-exact transition locations for |*δ*_*B*_| depend only on *α* and do not vary with waveform amplitude *A*_*n*_. The error in 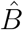 that will result in some samples failing bit-exact reconstruction is 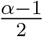 which corresponds to 0.72, 1.94, and 4.38 stream units for gains of 5, 10, and 20 mm/mV, respectively. However, 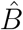 must be an integer, so we find the smallest integer exceeding this bound. Therefore, an error in 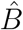 of 1, 2, and 5 stream units for gains of 5, 10, and 20 mm/mV, respectively, will result in some error in reconstruction, while smaller errors do not prevent bit-exact waveform reconstruction. Note that gains of 10 and 20 mm/mV have a “buffer” of 1 and 4 stream units, respectively, while a gain of 5 mm/mV does not, and even a 1 stream unit error in *B* at a gain of 5 mm/mV will cause failure of bit-exact reconstruction.

The bound where every waveform will have an error is 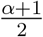 which corresponds to 1.72, 2.94, and 5.38 stream units for gains of 5, 10, and 20 mm/mV, respectively, which round up to 2, 3, and 6 stream units. This upper bound is exactly 1 stream unit greater than the lower bound.

### S.8. Expected Direct Stream Unit to mV Errors

We will assume that that errors between PDF reconstructed waveforms and the raw data in XML are only due to the rounding errors which are on the interval 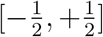. As discussed in detail in Supplement Section S.5, due to the double quantization of the waveforms, the rounding errors do not continuously fill the interval 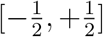. Instead, the values which are reachable are multiples of 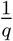 for integer *q*, where 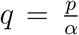. In all MUSE ECGs *q* = 25. Therefore, the errors take on equally spaced intervals with spacing of 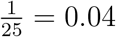, which gives us possible rounding errors of:

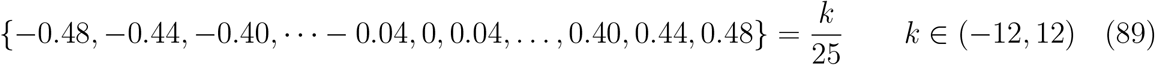

In this case, and assuming all rounding errors are approximately equally distributed, the MAE and RMSE (in stream units) associated with the rounding errors are therefore:

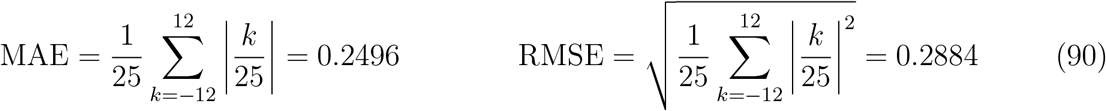

To convert stream units to *μ*V we simply use the conversion between stream units and *μ*V at a gain *G*:

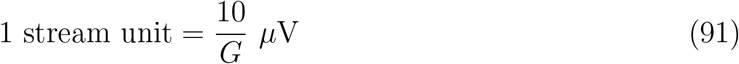

and the MAE and RMSE in *μ*V are:

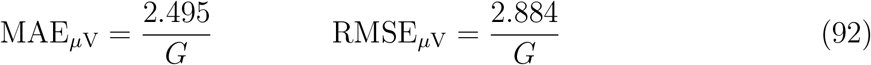

These values are extremely close to the observed MAE and RMSE in Table 2, with the minimal differences likely due to how there are not perfectly equal numbers of each possible quantized error, which is assumed in Eq. 90. The strong agreement between the observed MAE and RMSE with the predicted values further supports that the errors observed with direct stream unit to mV conversion are only due to rounding, and not some other systematic error/offset. This is further supported by the observed maximum errors for each gain which are exactly equal to the quantized limits of |0.5| stream units when converted to mV (see Table 2).

### S.9. Supplemental Tables

**Supplemental Table 1:** Definition of variables used throughout this manuscript.

| Variable | Description |
| --- | --- |
| $A$ | Integer ADU values recorded from ECG machine (values to be reconstructed) |
| $S$ | Integer PDF stream units plotted on the PDF |
| $Y$ | PDF stream units prior to rounding |
| $B$ | Integer offset used to position waveforms at specific locations on the PDF page |
| $V$ | Reconstructed waveform values in units of voltage |
| $\alpha$ | Number of stream units per 1 ADU |
| $G$ | ECG voltage gain in mm/mV |
| $T$ | ECG time gain in mm/sec (fixed at 25 mm/sec) |
| $g_v$ | Spacing of minor grid on voltage axis in stream units/mm |
| $g_t$ | Spacing of minor grid on time axis in stream units/mm |
| $m$ | Number of mV per 1 ADU (value not available from within PDF) |
| $b_v$ | Number of stream units per mV) |
| $b_t$ | Number of stream units per second) |

**Supplemental Table 2:** Summary of bit-exact waveform recovery for different PDF layouts and voltage gains in 2,336 ECGs from MUSE v9. Numbers represent number of leads with bit-exact reconstruction, the total number of leads processed (after excluding clipped leads), and the percent of the total. After excluding clipping, bit-exact recovery was present for 100% of leads in all formats and gains. *calcleads format does not reconstruct leads aVR, aVL, and aVF in any 3×4 format because these leads are not simultaneous with leads I and II, and as a result there are fewer total leads assessed in 3×4 in calcleads mode.

| PDF Layout | Lead Type | 10 mm/mV Gain |
| --- | --- | --- |
| $12 \times 1$ | calcleads* | 27879/27879<br>100% |
|  | pdfleads | 27953/27953<br>100% |
| $3 \times 4 + 3$ | calcleads* | 27994/27994<br>100% |
|  | pdfleads | 35003/35003<br>100% |

**Supplemental Table 3:**
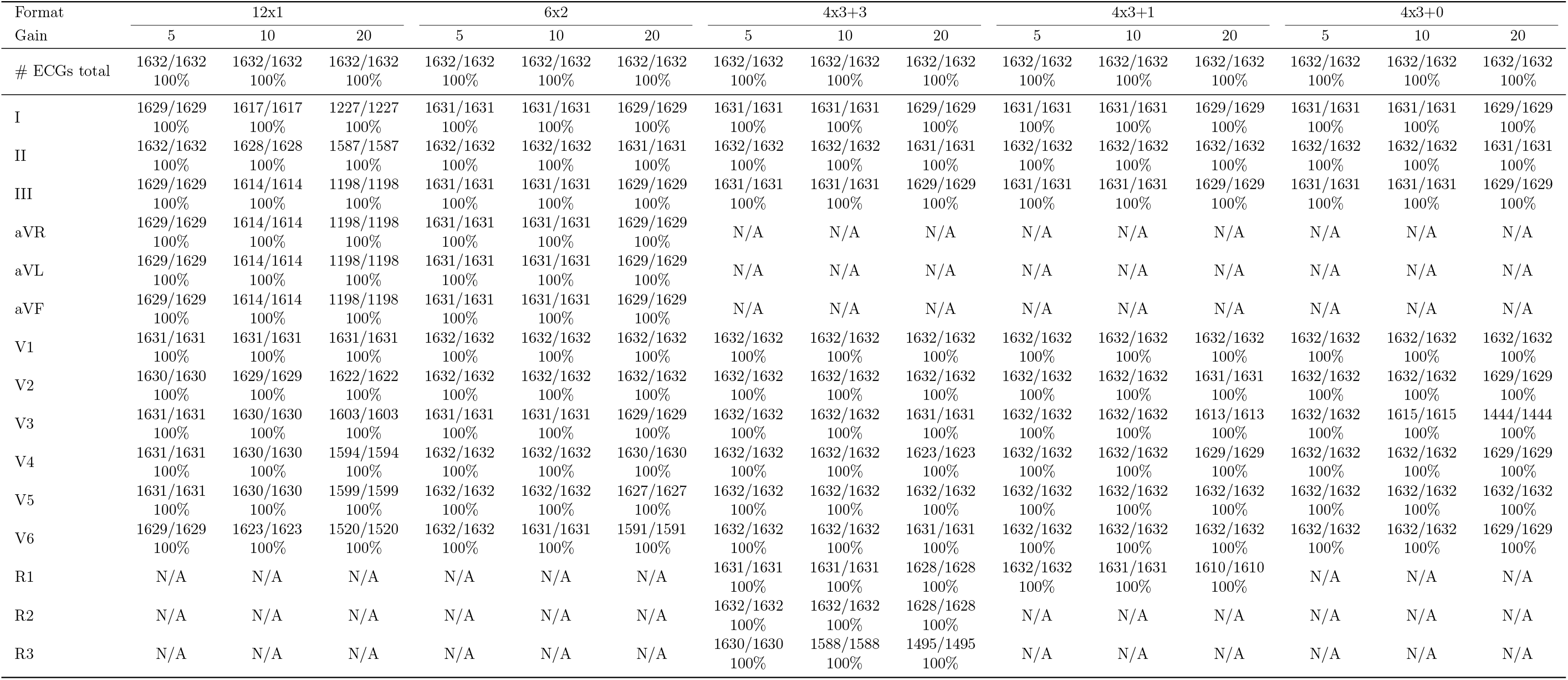
Per-lead results of bit-exact waveform recovery for different PDF layouts and voltage gains in 1,632 ECGs from MUSE v10 with waveforms reconstructed using calcleads mode. Numbers represent number of leads with bit-exact reconstruction, the total number of leads processed (after excluding clipped leads), and the percent of the total. After excluding clipping, bit-exact recovery was present for 100% of leads in all formats and gains. calcleads format does not reconstruct leads aVR, aVL, and aVF in any 3×4 format because these leads are not simultaneous with leads I and II, and as a result there are fewer total leads assessed in 3×4 formats. The R1, R2, and R3 rows represent the 3 possible rhythm strips which can be variable leads.

**Supplemental Table 4:** Per-lead results of bit-exact waveform recovery for different PDF layouts and voltage gains in 1,632 ECGs from MUSE v10 with waveforms reconstructed using pdfleads mode. Numbers represent number of leads with bit-exact reconstruction, the total number of leads processed (after excluding clipped leads), and the percent of the total. After excluding clipping, bit-exact recovery was present for 100% of leads in all formats and gains. he R1, R2, and R3 rows represent the 3 possible rhythm strips which can be variable leads.

| Format | 12x1 |  |  | 6x2 |  |  | 4x3+3 |  |  | 4x3+1 |  |  | 4x3+0 |  |  |
| --- | --- | --- | --- | --- | --- | --- | --- | --- | --- | --- | --- | --- | --- | --- | --- |
|  | 5 | 10 | 20 | 5 | 10 | 20 | 5 | 10 | 20 | 5 | 10 | 20 | 5 | 10 | 20 |
| # ECGs total | 1632/1632<br>100% | 1632/1632<br>100% | 1632/1632<br>100% | 1632/1632<br>100% | 1632/1632<br>100% | 1632/1632<br>100% | 1632/1632<br>100% | 1632/1632<br>100% | 1632/1632<br>100% | 1632/1632<br>100% | 1632/1632<br>100% | 1632/1632<br>100% | 1632/1632<br>100% | 1632/1632<br>100% | 1632/1632<br>100% |
| I | 1629/1629<br>100% | 1617/1617<br>100% | 1227/1227<br>100% | 1631/1631<br>100% | 1631/1631<br>100% | 1629/1629<br>100% | 1631/1631<br>100% | 1631/1631<br>100% | 1629/1629<br>100% | 1631/1631<br>100% | 1631/1631<br>100% | 1629/1629<br>100% | 1631/1631<br>100% | 1631/1631<br>100% | 1629/1629<br>100% |
| II | 1632/1632<br>100% | 1628/1628<br>100% | 1587/1587<br>100% | 1632/1632<br>100% | 1632/1632<br>100% | 1631/1631<br>100% | 1632/1632<br>100% | 1632/1632<br>100% | 1631/1631<br>100% | 1632/1632<br>100% | 1632/1632<br>100% | 1632/1632<br>100% | 1632/1632<br>100% | 1632/1632<br>100% | 1631/1631<br>100% |
| III | 1631/1631<br>100% | 1630/1630<br>100% | 1625/1625<br>100% | 1631/1631<br>100% | 1631/1631<br>100% | 1630/1630<br>100% | 1631/1631<br>100% | 1631/1631<br>100% | 1630/1630<br>100% | 1631/1631<br>100% | 1631/1631<br>100% | 1629/1629<br>100% | 1631/1631<br>100% | 1630/1630<br>100% | 1586/1586<br>100% |
| aVR | 1631/1631<br>100% | 1631/1631<br>100% | 1630/1630<br>100% | 1631/1631<br>100% | 1631/1631<br>100% | 1630/1630<br>100% | 1632/1632<br>100% | 1632/1632<br>100% | 1631/1631<br>100% | 1632/1632<br>100% | 1632/1632<br>100% | 1631/1631<br>100% | 1632/1632<br>100% | 1632/1632<br>100% | 1631/1631<br>100% |
| aVL | 1631/1631<br>100% | 1631/1631<br>100% | 1630/1630<br>100% | 1631/1631<br>100% | 1630/1630<br>100% | 1629/1629<br>100% | 1632/1632<br>100% | 1632/1632<br>100% | 1631/1631<br>100% | 1632/1632<br>100% | 1632/1632<br>100% | 1631/1631<br>100% | 1632/1632<br>100% | 1632/1632<br>100% | 1631/1631<br>100% |
| aVF | 1631/1631<br>100% | 1631/1631<br>100% | 1631/1631<br>100% | 1631/1631<br>100% | 1626/1626<br>100% | 1541/1541<br>100% | 1632/1632<br>100% | 1632/1632<br>100% | 1632/1632<br>100% | 1632/1632<br>100% | 1632/1632<br>100% | 1632/1632<br>100% | 1632/1632<br>100% | 1632/1632<br>100% | 1621/1621<br>100% |
| V1 | 1631/1631<br>100% | 1631/1631<br>100% | 1631/1631<br>100% | 1632/1632<br>100% | 1632/1632<br>100% | 1632/1632<br>100% | 1632/1632<br>100% | 1632/1632<br>100% | 1632/1632<br>100% | 1632/1632<br>100% | 1632/1632<br>100% | 1632/1632<br>100% | 1632/1632<br>100% | 1632/1632<br>100% | 1632/1632<br>100% |
| V2 | 1630/1630<br>100% | 1629/1629<br>100% | 1622/1622<br>100% | 1632/1632<br>100% | 1632/1632<br>100% | 1632/1632<br>100% | 1632/1632<br>100% | 1632/1632<br>100% | 1632/1632<br>100% | 1632/1632<br>100% | 1632/1632<br>100% | 1631/1631<br>100% | 1632/1632<br>100% | 1632/1632<br>100% | 1629/1629<br>100% |
| V3 | 1631/1631<br>100% | 1630/1630<br>100% | 1603/1603<br>100% | 1631/1631<br>100% | 1631/1631<br>100% | 1629/1629<br>100% | 1632/1632<br>100% | 1632/1632<br>100% | 1631/1631<br>100% | 1632/1632<br>100% | 1632/1632<br>100% | 1613/1613<br>100% | 1632/1632<br>100% | 1615/1615<br>100% | 1444/1444<br>100% |
| V4 | 1631/1631<br>100% | 1630/1630<br>100% | 1594/1594<br>100% | 1632/1632<br>100% | 1632/1632<br>100% | 1630/1630<br>100% | 1632/1632<br>100% | 1632/1632<br>100% | 1623/1623<br>100% | 1632/1632<br>100% | 1632/1632<br>100% | 1629/1629<br>100% | 1632/1632<br>100% | 1632/1632<br>100% | 1629/1629<br>100% |
| V5 | 1631/1631<br>100% | 1630/1630<br>100% | 1599/1599<br>100% | 1632/1632<br>100% | 1632/1632<br>100% | 1627/1627<br>100% | 1632/1632<br>100% | 1632/1632<br>100% | 1632/1632<br>100% | 1632/1632<br>100% | 1632/1632<br>100% | 1632/1632<br>100% | 1632/1632<br>100% | 1632/1632<br>100% | 1632/1632<br>100% |
| V6 | 1629/1629<br>100% | 1623/1623<br>100% | 1520/1520<br>100% | 1632/1632<br>100% | 1631/1631<br>100% | 1591/1591<br>100% | 1632/1632<br>100% | 1632/1632<br>100% | 1631/1631<br>100% | 1632/1632<br>100% | 1632/1632<br>100% | 1632/1632<br>100% | 1632/1632<br>100% | 1632/1632<br>100% | 1629/1629<br>100% |
| R1 | N/A | N/A | N/A | N/A | N/A | N/A | 1631/1631<br>100% | 1631/1631<br>100% | 1628/1628<br>100% | 1632/1632<br>100% | 1631/1631<br>100% | 1610/1610<br>100% | N/A | N/A | N/A |
| R2 | N/A | N/A | N/A | N/A | N/A | N/A | 1632/1632<br>100% | 1632/1632<br>100% | 1628/1628<br>100% | N/A | N/A | N/A | N/A | N/A | N/A |
| R3 | N/A | N/A | N/A | N/A | N/A | N/A | 1630/1630<br>100% | 1588/1588<br>100% | 1495/1495<br>100% | N/A | N/A | N/A | N/A | N/A | N/A |

**Supplemental Table 5:**
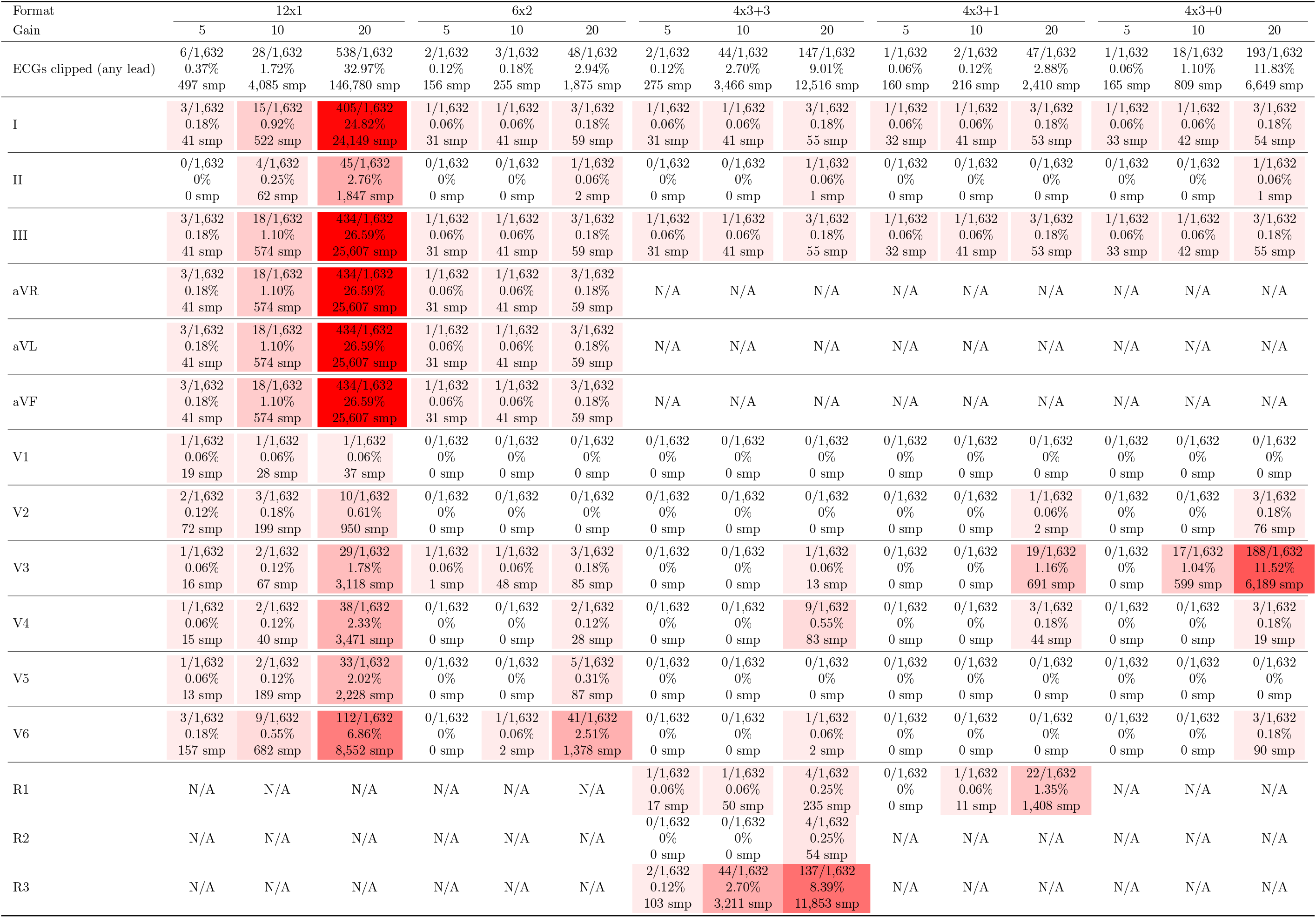
Per-lead clipping results for different PDF layouts and voltage gains in 1,632 ECGs from MUSE v10 with waveforms reconstructed using calcleads mode. The red shading indicates the relative percent of leads with clipping; darker red indicates more leads clipped. In calcleads mode leads III, aVR, aVL, and aVF are calculated from leads I and II, so any clipping in leads I and II is inherited by the calculated leads to avoid inaccurate reconstruction of the calcualted leads. The R1, R2, and R3 rows represent the 3 possible rhythm strips which can be variable leads.

**Supplemental Table 6:**
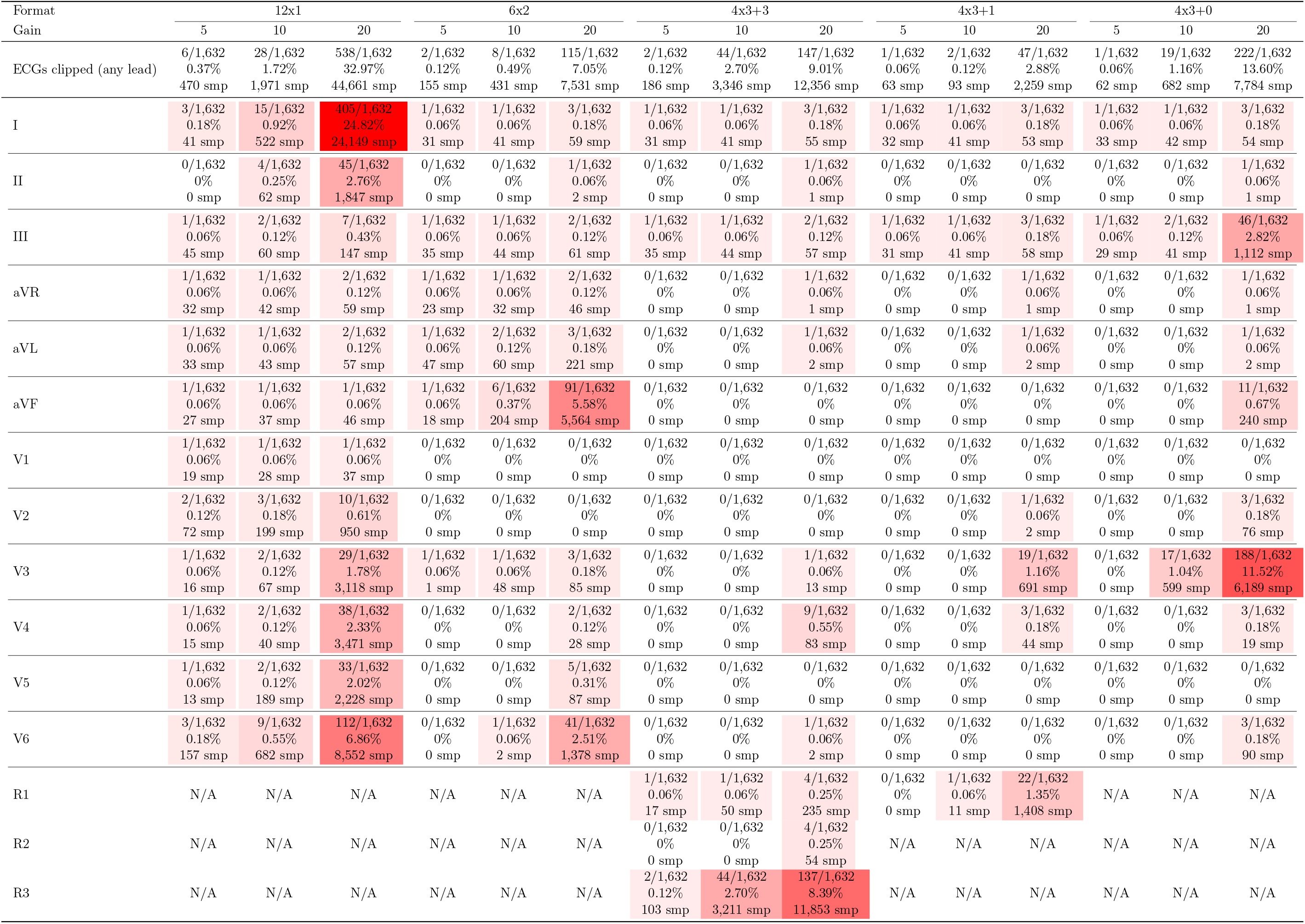
Per-lead clipping results for different PDF layouts and voltage gains in 1,632 ECGs from MUSE v10 with waveforms reconstructed using pdfleads mode. The red shading indicates the relative percent of leads with clipping; darker red indicates more leads clipped. The R1, R2, and R3 rows represent the 3 possible rhythm strips which can be variable leads.

**Supplemental Table 7:** Per-lead reconstruction errors associated with clipping in 28 ECGs in 12 × 1 format at 10 mm/mV gain. Note how the most frequent clipping locations are leads I and V6 which are at the top and bottom of the ECG page.

| Lead | N | Max Error ($\mu\text{V}$) | MAE ($\mu\text{V}$) | RMSE ($\mu\text{V}$) | Correlation $r$ |
| --- | --- | --- | --- | --- | --- |
| I | 15 | $2236 \pm 8153$ | $1.503 \pm 4.272$ | $109.6 \pm 403.4$ | $0.9687 \pm 0.0976$ |
| II | 4 | $146 \pm 141$ | $0.2462 \pm 0.183$ | $4.768 \pm 3.691$ | $0.9977 \pm 0.0033$ |
| III | 2 | $1.59 \times 10^4 \pm 2.21 \times 10^4$ | $20.8 \pm 29.12$ | $811.9 \pm 1139$ | $0.9154 \pm 0.1154$ |
| aVR | 1 | $1.49 \times 10^4$ | 15.76 | 735.9 | 0.8713 |
| aVL | 1 | $3.04 \times 10^4$ | 26.93 | 1585 | 0.7092 |
| aVF | 1 | $1.43 \times 10^4$ | 10.74 | 724.5 | 0.8260 |
| V1 | 1 | 8696 | 26.66 | 426.9 | 0.87439 |
| V2 | 3 | $4479 \pm 4103$ | $21.44 \pm 18.27$ | $276.3 \pm 224.9$ | $0.9217 \pm 0.0673$ |
| V3 | 2 | $4964 \pm 4640$ | $15.47 \pm 12.37$ | $240.7 \pm 224.2$ | $0.9425 \pm 0.0669$ |
| V4 | 2 | $4268 \pm 5151$ | $11.72 \pm 15.3$ | $199.1 \pm 254.6$ | $0.9498 \pm 0.0617$ |
| V5 | 2 | $3844 \pm 5083$ | $13.07 \pm 10.25$ | $193.4 \pm 223.8$ | $0.9530 \pm 0.0536$ |
| V6 | 9 | $934.9 \pm 2310$ | $4.475 \pm 7.716$ | $50.77 \pm 108.2$ | $0.9840 \pm 0.0268$ |

### S.10. Supplemental Figures

**Supplemental Figure 1:**
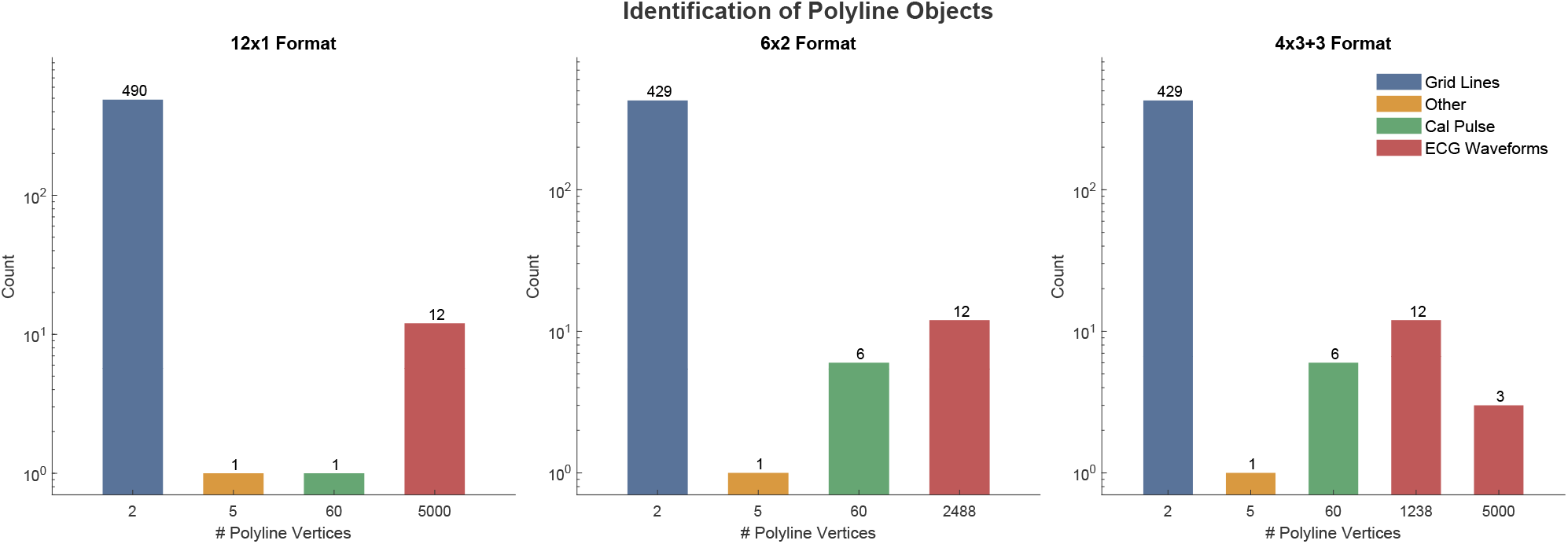
Identification and distribution of polylines with different numbers of vertices.

**Supplemental Figure 2:**
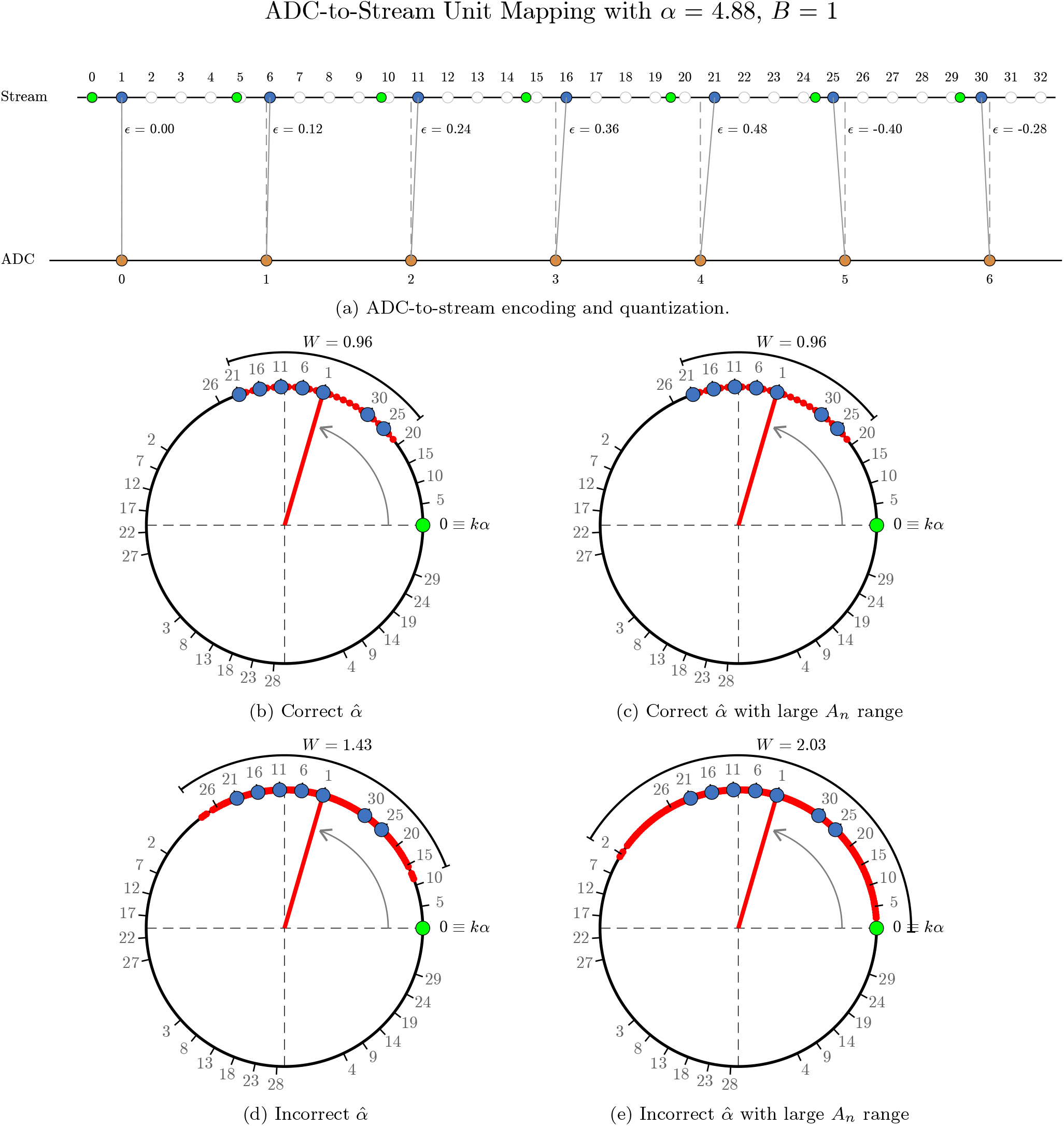
Illustration of taking modulo 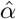 of monotonically increasing stream units. This is geometrically equivalent to wrapping the stream units counterclockwise around a circle of radius 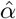. Note that all points in stream units that are integer multiples of 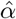 end up at the “0” value denoted by a green dot on the number line and on the circle. The values on the stream unit number line (blue dots) are mapped to their equivalent numbered position on the circle as the values are wrapped around. The blue points, which are stream values that are obtained from ADU values all cluster in the arc denoted in red. Other values that are not occupied span the full circumference of the circle. **Panels B and C:** illustrate that if there is no error in 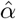, the residue arc ≤0.96 stream units regardless of signal amplitude. **Panel D:** If 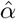 is incorrect, the residue width is *>* 0.96 stream units. **Panel E:** If 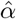 is incorrect, the residue width depends on the amplitude of the ADU values that are encoded; As the range of *A*_*n*_ increases, the arc width increases.

**Supplemental Figure 3:**
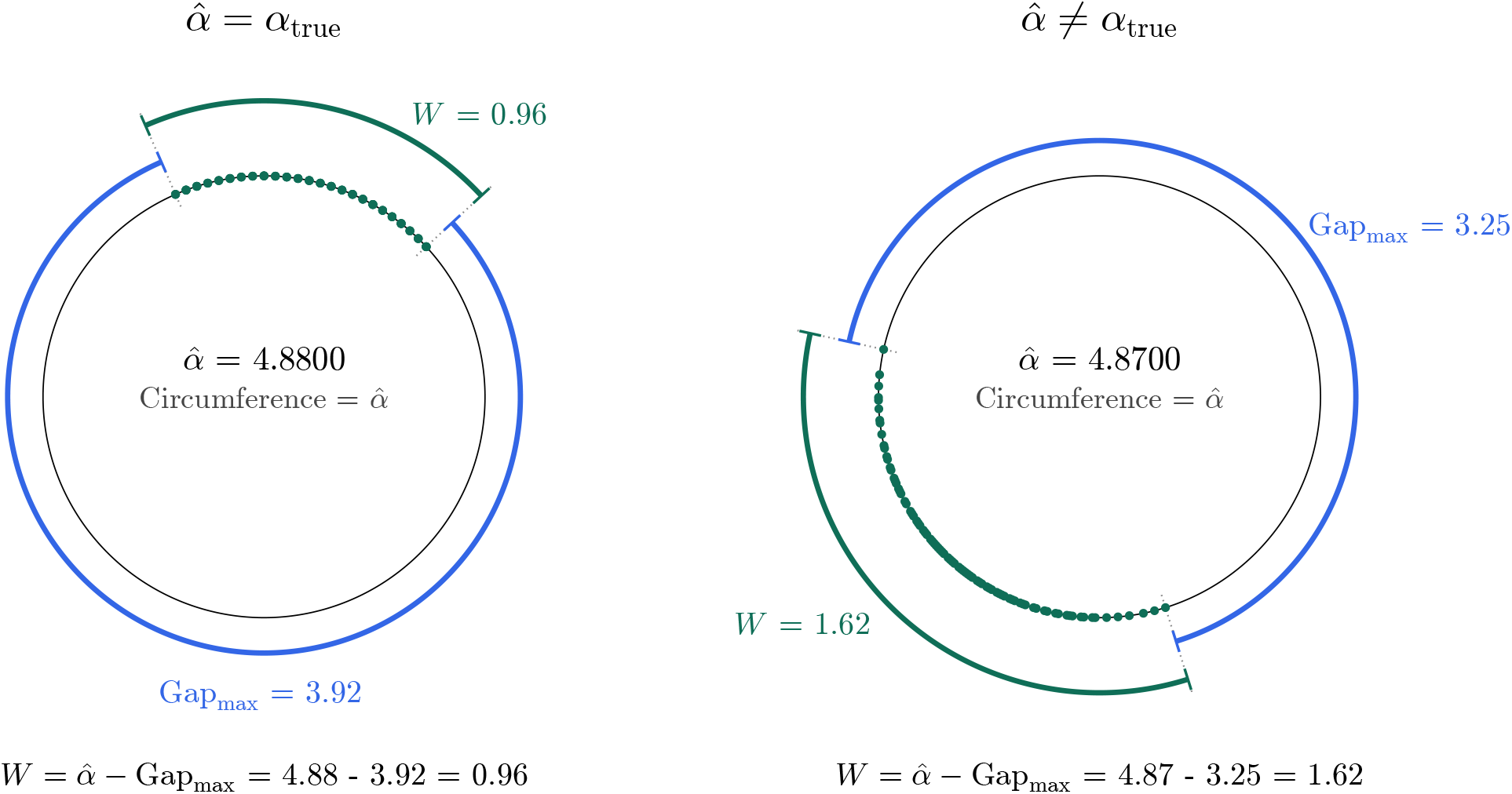
Calculating the width of residues (in green) by subtracting the maximum gap from the circle circumference. As the residues live on a circle, standard subtraction is insufficient to determine gaps between residues.

**Supplemental Figure 4:**
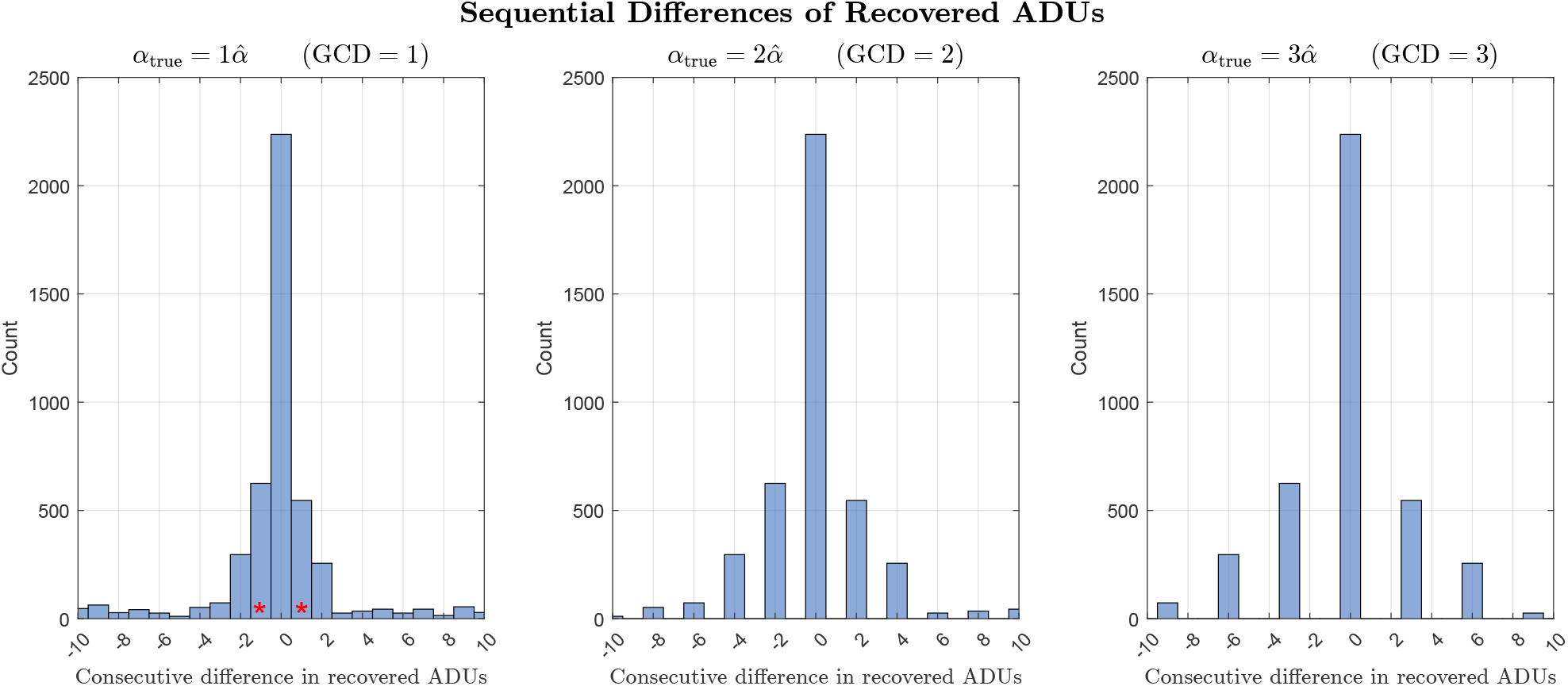
Histogram of sequential, non-zero differences in Reconstructed ADUs. When 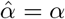 (left panel), the greatest common divisor (GCD) is 1 (see red asterisks). For any integer submultiple of *α*, the GCD is *>* 1.

**Supplemental Figure 5:**
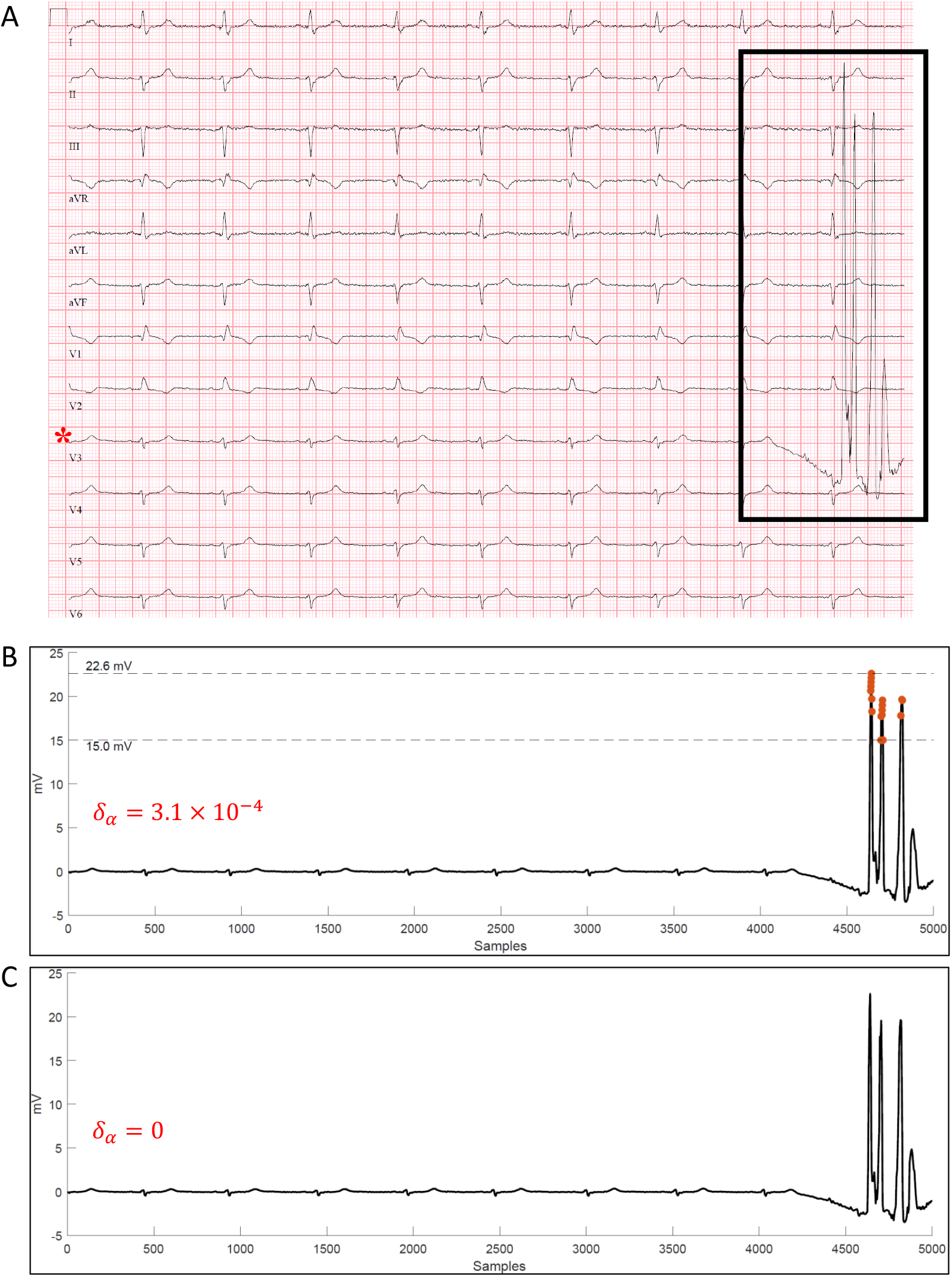
Single ECG lead (V3) with a non-physiological artifact which due to its 5 mm/mV gain (Panel A), location, and voltage magnitude had 17 samples (orange points) that were off by exactly 1 ADU due to an insignificant number of significant digits in 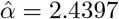 instead of 2.44 (an error of 3.1 × 10^−4^) (Panel B). The error occurred only for points that were 15.0–22.6 mV in amplitude. Note that based on Supplement Section S.6, the minimum error where failure of bit-exact reconstruction begins to appear is given as 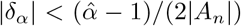. Using |*δ*_*α*_| = 3.1 × 10^−4^ and 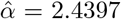, the minimum value of *A*_*n*_ where bit-exact reconstruction breaks down is 11.5 mV (see Supplemental Figure 6). When the correct 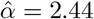 is used, (Panel C) every point has bit-exact reconstruction

**Supplemental Figure 6:**
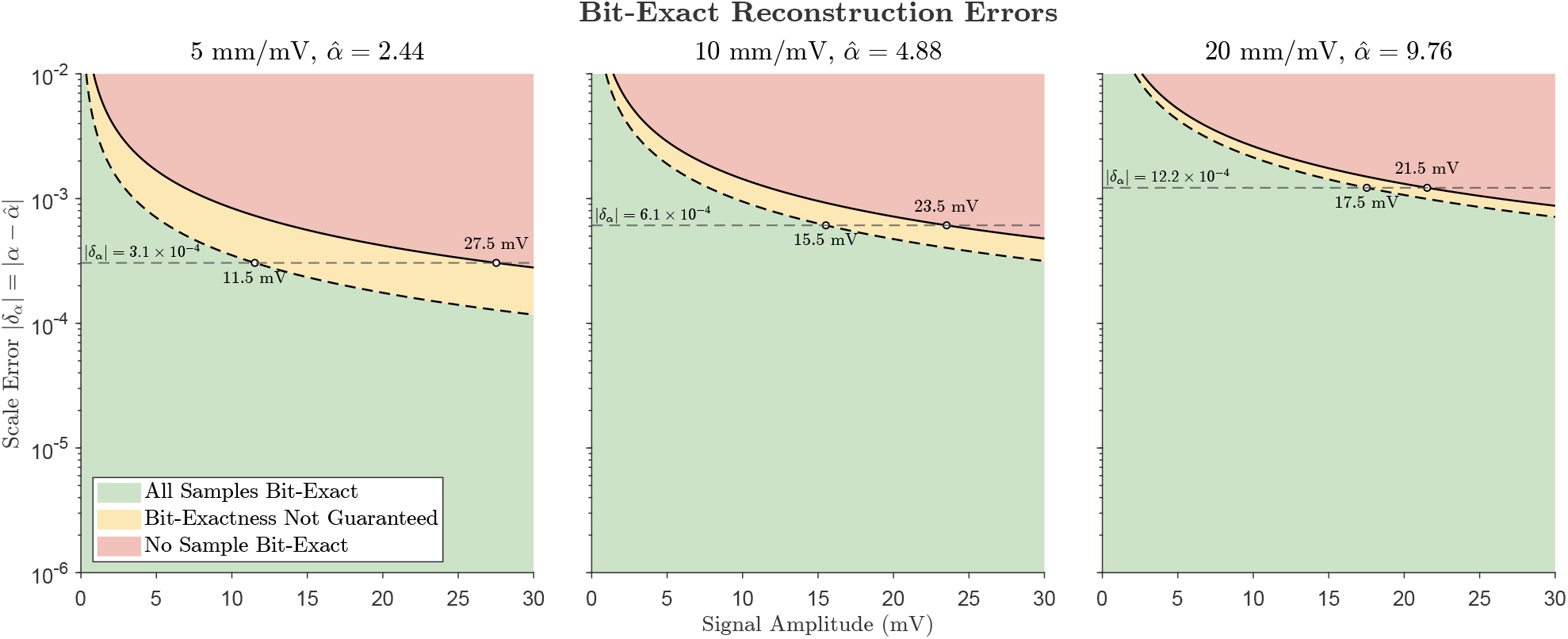
Bit-exact reconstruction as a function of signal amplitude and error in the assumed stream-to-ADU relationship 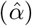, at gains of 5 mm/mV 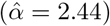, 10 mm/mV 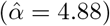, and 20 mm/mV 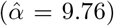. Because the rounding error depends on the product *δ*_*α*_*A*_*n*_, scale error and a signal amplitude trade off directly. Below the first transition (dashed line, 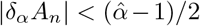) in the green shaded area, every sample is recovered exactly regardless of the value of *ϵ*_*n*_. Between the two transitions, in yellow, only samples whose rounding residual lies near 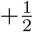 cross an ADU rounding boundary, so failures are sparse and confined to the extremes of the waveform while the remainder is recovered exactly. In this area, bit-exactness is no longer guaranteed but is still frequently achieved. Above the second transition (solid line, 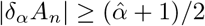), no value of *ϵ*_*n*_ can prevent failure and every sample at that amplitude is reconstructed in error, although lower-amplitude samples within the same lead remain exact. The dotted horizontal line marks the an example of a small error in scale 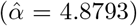 used before 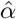 was determined in exact rational form. At a gain of 5 mm/mV, *δ*_*α*_ = 3.05 ×10^−4^ and transitions fall at 11.5 and 27.5 mV. Under the exact rational scale *δ*_*α*_ = 0 and reconstruction is bit-exact at any amplitude. See Supplemental Figure 5 for additional information

**Supplemental Figure 7:**
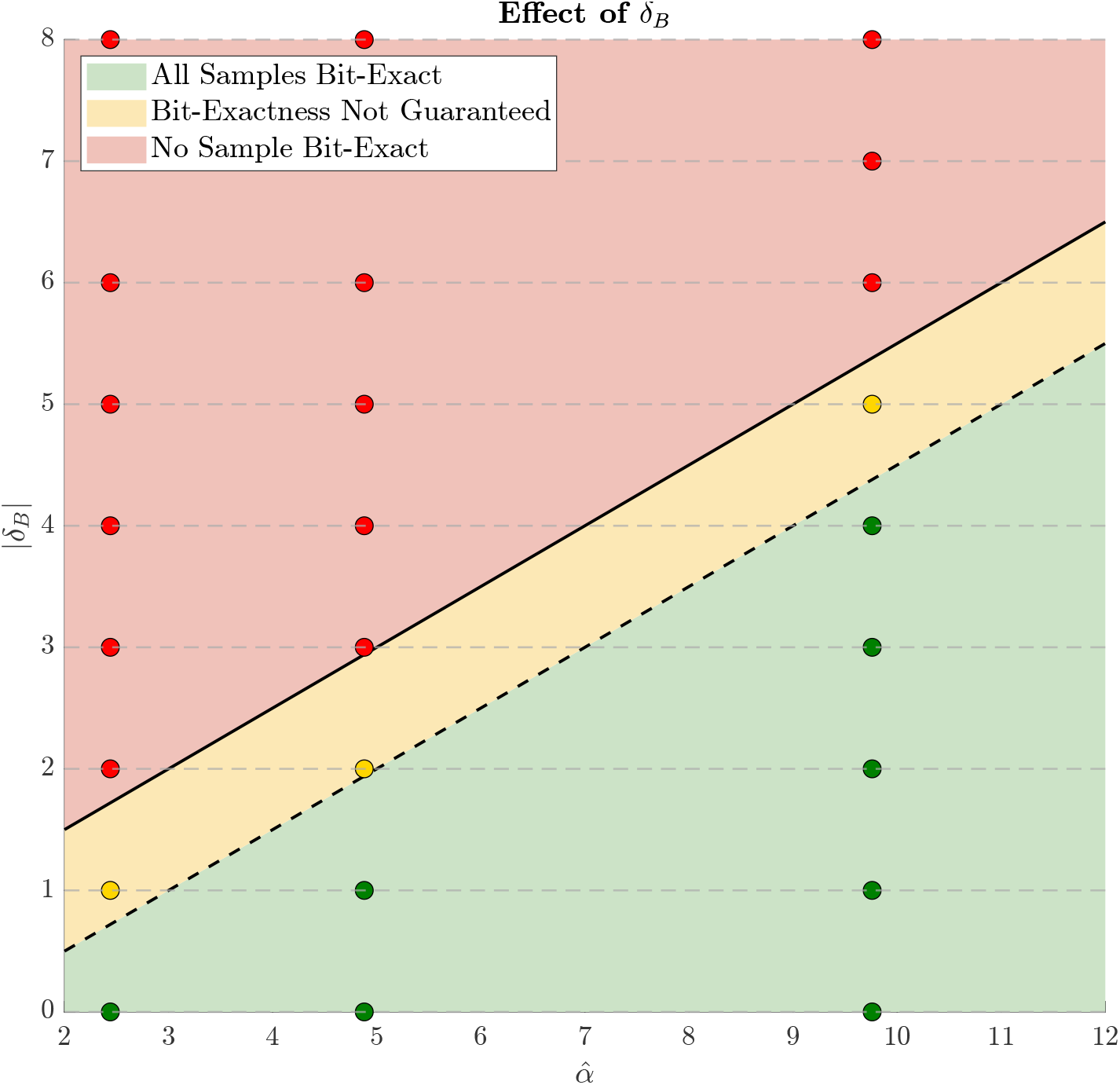
Bit-exact reconstruction as a function of error in the assumed baseline offset *δ*_*B*_, at gains of 5 mm/mV 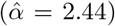, 10 mm/mV 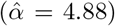, and 20 mm/mV 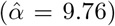. *δ*_*B*_ is an integer, as there are no fractional stream units. In the green shaded area, *δ*_*B*_ is not large enough to impact bit-exact reconstruction. In the yellow shaded area bit-exact reconstruction can no longer be confirmed, but every sample will not be corrupted. In the red shaded area, every sample will have an error. At a gain of 5 mm/mV 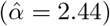 there is no margin for errors in the value of *B*, as even if *δ*_*B*_ = 1 we end up in the yellow area. At a gain of 10 mm/mV 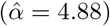 there is a 1 stream unit safety margin, and at 20 mm/mV 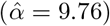 there is a 4 stream unit safety margin before reaching the yellow area. Notably, the baseline residue test will fail if *δ*_*B*_ ≠ 0 even if bit-exact reconstruction is still possible (for *δ*_*B*_ that area in the green area).

**Supplemental Figure 8:**
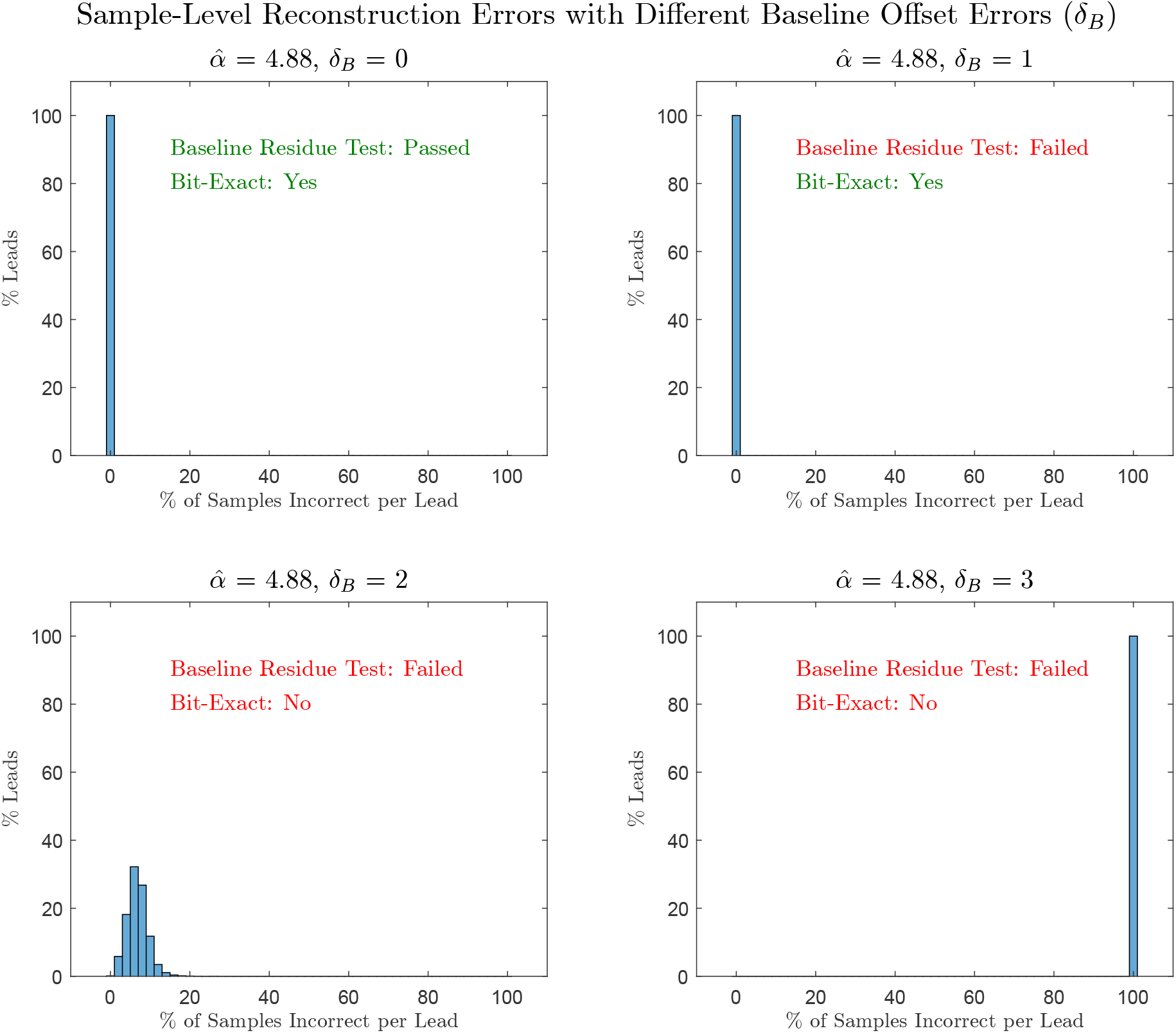
Sample level reconstruction errors at different values of offset errors (*δ*_*B*_) at a gain of 10 mm/mV (*α* = 4.88). The transition points for bit-exact certification are ⌈(*α* ± 1)/2⌉, which correspond to 2 and 3 stream units. When *δ*_*B*_ = 0 no samples are incorrect, the baseline residue test passes, and reconstruction is bit-exact. If *δ*_*B*_ = 1, the baseline residue test fails, but the error is not enough to cross any rounding thresholds (⌈(*α* − 1)/2⌉), and as a result, reconstruction is still bit-exact. Once *δ*_*B*_ = 2, the error exceeds the lower limit of ⌈(*α* − 1)/2⌉, and as a result, in addition to failing certification, samples begin to lose bit-exactness depending on their rounding errors. For *δ*_*B*_ = 3, the error is large enough and above the upper bound ⌈(*α* + 1)/2⌉, so every sample loses bit-exactness.

